# Clinical hypoxemia score for outpatient child pneumonia care lacking pulse oximetry in Africa and South Asia

**DOI:** 10.1101/2023.02.25.23286448

**Authors:** Holly B. Schuh, Shubhada Hooli, Salahuddin Ahmed, Carina King, Arunangshu D. Roy, Norman Lufesi, ASMD Ashraful Islam, Tisungane Mvalo, Nabidul H. Chowdhury, Amy Sarah Ginsburg, Tim Colbourn, William Checkley, Abdullah H. Baqui, Eric D. McCollum

## Abstract

**Background:** Pulse oximeters are not routinely available in outpatient clinics in low- and middle-income countries. We derived clinical scores to identify hypoxemic child pneumonia.

**Methods:** This was a retrospective pooled analysis of two outpatient datasets of 3-35 month olds with World Health Organization (WHO)-defined pneumonia in Bangladesh and Malawi. We constructed, internally validated, and compared fit & discrimination of four models predicting SpO2<93% and <90%: (1) Integrated Management of Childhood Illness guidelines, (2) WHO-composite guidelines, (3) Independent variable least absolute shrinkage and selection operator (LASSO); (4) Composite variable LASSO. Results: 12,712 observations were included. The independent and composite LASSO models discriminated moderately (both C-statistic 0.77) between children with a SpO2<93% and ≥94%; model predictive capacities remained moderate after adjusting for potential overfitting (C-statistic 0.74 and 0.75). The IMCI and WHO-composite models had poorer discrimination (C-statistic 0.56 and 0.68) and identified 20.6% and 56.8% of SpO2<93% cases. The highest score stratum of the independent and composite LASSO models identified 46.7% and 49.0% of SpO2<93% cases. Both LASSO models had similar performance for a SpO2<90%.

**Conclusions:** In the absence of pulse oximeters, both LASSO models better identified outpatient hypoxemic pneumonia cases than the WHO guidelines. Score external validation and implementation are needed.

## Introduction

The burden of child pneumonia mortality predominantly occurs in low-income and middle-income countries (LMICs).^1^ Hypoxemia – a low blood oxyhemoglobin saturation – conveys increased pneumonia mortality risk, yet most children in LMICs lack pulse oximeter access.^2,3^ Pulse oximeters identify hypoxemia by non-invasively measuring the peripheral arterial oxyhemoglobin saturation (SpO_2_).^4^ To simplify diagnosis in LMICs the World Health Organization (WHO) Integrated Management of Childhood Illness (IMCI) guidelines consider pneumonia a clinical syndrome.^5^ Although IMCI recommends oximeter use when available, it also provides guidance for settings without pulse oximetry.^5^ However, when implemented without pulse oximeters, IMCI missed almost 70% of outpatient pneumonia cases with a SpO_2_<90% in Malawi and nearly 90% in Bangladesh.^6,7^ As most children first access health systems at outpatient clinics, improving outpatient hypoxemia identification may be key to reducing LMIC pneumonia mortality.^4^

Prior research attempted to determine whether clinical signs accurately identify a SpO_2_<90% in hospitalized children.^8,9^ While research showed high specificity of clinical signs for a SpO_2_ <90%, sensitivity was low.^8,9^ Additionally, recent data showed elevated mortality among children with a SpO_2_ 90-92% than higher SpO_2_ levels.^7,10-12^ These findings challenge the currently recommended SpO_2_<90% threshold for hospitalization and oxygen treatment in LMICs.

We sought to utilize two unique, contemporary outpatient pediatric pneumonia datasets from Bangladesh and Malawi to accomplish three objectives.^13,14^ First, evaluate IMCI performance for identifying hypoxemia at a higher SpO_2_ threshold (<93%) than recommended. Second, examine whether other combinations of clinical features better identify children with a SpO_2_<93%, followed by development and internal validation of hypoxemia clinical scores feasible for LMICs. Third, repeat these analyses using the currently recommended SpO_2_<90% threshold.

## METHODS

### Settings

We used data from Malawi and Bangladesh. Malawi is an African country with an under 5 mortality rate of 49/1,000 births.^15^ This study included 18 clinics in Mchinji and Lilongwe districts with a 1.2 million population catchment area, at 1,000-1,100 meters altitude.^13^ From October 2011–June 2014, non-physician clinicians and nurses at clinics were IMCI trained and documented care of 0-59 month olds with WHO-defined pneumonia.^13^ Providers used Acare® pulse oximeters with adult clip probes on the big toe if <2 years old or weighing <10kg.^6^ Training, data collection, and supervision methodology has been published.^13^

Bangladesh has an under 5 mortality rate of 3/1,000 live births.^16^ Since 2001, Projahnmo, a partnership of Johns Hopkins University with the Government of Bangladesh’s Ministry of Health and Family Welfare, International Centre for Diarrhoeal Disease Research, Bangladesh, Shimantik, and the Child Health Research Foundation conducted community-based surveillance in Zakiganj subdistrict of Sylhet district in Bangladesh.^14^ From May 2015–September 2017 Projahnmo expanded surveillance into two additional subdistricts (Kanaighat, Beanibazar) and augmented IMCI clinic care of three government Upazila Health Complexes.^14^ Altogether these subdistricts have a 770,000 population at 17-23 meters altitude. Upazila Health Complexes provide outpatient and emergency care and limited inpatient pediatric services. IMCI clinic care was provided by Projahnmo physicians per IMCI guidelines.^14^ From October 2017 Projahnmo physicians measured the SpO_2_ of 3-35 month olds with suspected pneumonia using a Masimo Rad-5® pulse oximeter with a LNCS® Y-I wrap sensor on the big toe. Parent study methodology is published.^14^

### Inclusion and Exclusion Criteria

We generated an analytic sample of healthcare visits from Bangladesh and Malawi datasets. Inclusion criteria were: valid SpO_2_, IMCI-defined non-severe or severe pneumonia (2014 guidelines),^5^ and age 3-35 months, as the Bangladesh study population with a SpO_2_ was limited to this age.^17^ See Supplemental Material for study definitions. Implicit to this analysis we assumed SpO_2_ was unavailable and excluded it from pneumonia definitions.

### Variables

Our primary outcome was a SpO_2_<93%; SpO_2_<90% was secondary. We explored associations between clinical variables and SpO_2_ ranges to evaluate <93% as the primary outcome. We selected variables *a priori*: sex, age, weight-for-age z-score (WAZ), chest indrawing, wheezing, severe respiratory distress (grunting, head nodding, nasal flaring, and/or age-adjusted very fast breathing), cyanosis, fever (temperature ≥ 38°C), and WHO-defined general danger signs (stridor, inability to feed/drink, convulsions, and/or lethargy).^8,18^

### Analysis

We evaluated missingness using a 5% threshold. We used Chi squared and Fisher’s exact tests for proportions, Wilcoxon rank-sum for non-parametric data, and Student’s t-test for normally distributed data comparisons. We reviewed individual-level variables and their associated SpO_2_<93% and <90% predictive quality. For SpO_2_<93% and <90% model development we randomly split the dataset into derivation (70%) and validation (30%) sets balanced by outcome and country. For each model we fit a logistic regression model with hypoxemia as the binary outcome measure. We allowed selection of *country* as a fixed effect to account for significant differences by country. Thereafter, we used a random intercept in each post selection model to control for country after interrogating the suitability of this approach with the Hausman test.^19^ All analyses were by Stata 16.1 (StataCorp, College Station, TX).

### Model development

*IMCI guidelines (IMCI model) and Composite WHO guidelines (WHO-composite model)* The IMCI model reflects 2014 IMCI referral criteria (Supplemental Material).^5^ The WHO-composite model is a composite of four WHO guidelines.^5,18,20,21^ We fit both models using the *logistic* command for implementing multivariable, maximum-likelihood logit models to obtain odds ratios (and 95% confidence intervals) comparing the odds of hypoxemia versus non-hypoxemia.

### Independent variable LASSO model (Independent LASSO model) and Composite variable LASSO model (Composite LASSO model)

We used the least absolute shrinkage and selection operator (LASSO) reduction method, testing two selection modes ((1) 10-fold cross-validation selection and (2) adaptive selection) using the derivation dataset to develop both models from an expanded variable list of the IMCI and WHO-composite models (Supplemental Material).^22,23^ For Independent LASSO we used *singular* variables (i.e., *independent*) whereas for Composite LASSO we used *composite* variables for ‘danger signs’ (WHO-defined general danger signs) and ‘severe respiratory distress.’ For both models we compared the two methods using the C-statistic based on predicted estimates, sensitivity, and specificity. If we found no statistical difference between methods, we used the selection results from the simplest model to implement an unsupervised approach (i.e. selection of the full variable rather than one category of a three-category variable) to refit both models.

### Hypoxemia score development and validation

We compared discriminatory power and model fit (C-statistic and Bayesian Information Criteria (BIC)) of the four maximum-likelihood logit models using the derivation dataset models.^24,25^ Using an unsupervised approach (i.e., if only two age groups were selected, we retained all age categories), each of the LASSO model covariates were kept on the log scale, rounded to the nearest 0.5, and doubled to form an integer.^10,26,27^ We then split the score into approximately equally sized quintiles to create hypoxemia risk categories.

Using the validation dataset, LASSO model scores were estimated by child, and score discriminatory power to identify children with and without hypoxemia was determined. Scores were not developed using IMCI and WHO-composite models. The C-statistic, sensitivity, specificity, positive and negative predictive value (PPV and NPV), and positive and negative likelihood ratios (LR+ and LR-) were compared across all models. We adjusted C-statistics for optimism by bootstrapping (200 repetitions) to account for any overfitting.^28^ A C-statistic 0.71-0.80 was considered moderate and >0.80 as excellent discriminatory power.^29^ We applied the same analysis methodology for SpO_2_<90%.

### Ethical Approval

Ethical approval was provided by National Health Sciences Research Committee of Malawi, the Ethics Committee of University College London, and The Bangladesh Medical Research Counsel, International Centre for Diarrhoeal Diseases Research, Johns Hopkins Bloomberg School of Public Health and School of Medicine Institutional Review Boards. Caregiver written consent was required in Bangladesh but not Malawi.

## RESULTS

### Study Population

We included 12,712 pneumonia cases; 63.6% (n=8,081) were from Bangladesh (Supplemental Material). Table 1 shows all participant characteristics by SpO_2_. A SpO_2_<93% was in 10.4% (1,328/12,712) of cases and 63.6% (845/1,328) of SpO_2_<93% cases were in Malawi. Most SpO_2_<93% cases had non-severe pneumonia (1,065/1,328; 81.4%) and, without a SpO_2_ measurement, were hospitalization ineligible per IMCI guidelines. A larger proportion of severe (263/1,198, 21.9%) than non-severe (1,065/11,514, 9.2%) cases had a SpO_2_<93%. A SpO_2_<90% was in 4.6% (602/12,712) of cases and in 3.8% (434/11,514) with non-severe disease. 2014 IMCI hospital referral criteria missed 72.0% (434/602) of SpO_2_<90% cases. While Bangladesh and Malawi case characteristics differed in frequency, other than WAZ<-3 and danger signs, they had similar crude associations with a SpO_2_<93% (Supplemental Material).

**Table 1.** Patient characteristics by SpO_2_<93% (full dataset, N=12,712)

|  | Total | SpO<sub>2</sub> ≥93% | SpO<sub>2</sub><93% | p-value |
| --- | --- | --- | --- | --- |
| Characteristics | N=12,712 | N=11,384<br>(89.6%) | N=1,328<br>(10.4%) |  |
| Study country, n (%) |  |  |  | <0.001 |
| Bangladesh | 8,081 (63.6%) | 7,598 (66.7%) | 483 (36.4%) |  |
| Malawi | 4,631 (36.4%) | 3,786 (33.3%) | 845 (63.6%) |  |
| Child age (months), median (IQR) | 12 (6, 19) | 12 (6, 20) | 10 (6, 17) | <0.001 |
| Child sex, n (%) |  |  |  | 0.36 |
| Male | 6,895 (54.2%) | 6,207 (54.5%) | 688 (51.8%) |  |
| Female | 5,530 (43.5%) | 4,950 (43.5%) | 580 (43.7%) |  |
| Missing | 287 (2.3%) | 227 (2.0%) | 60 (4.5%) |  |
| Weight for age z-score, n (%) |  |  |  | 0.056 |
| -2.0 | 9,441 (74.3%) | 8,463 (74.3%) | 978 (73.6%) |  |
| -3.0 ≤ z-score < -2.0 | 2,080 (16.4%) | 1,899 (16.7%) | 181 (13.6%) |  |
| < -3.0 | 852 (6.7%) | 760 (6.7%) | 92 (6.9%) |  |
| Missing | 339 (2.7%) | 262 (2.3%) | 77 (5.8%) |  |
| WHO danger signs, n (%) | 364 (2.9%) | 183 (1.6%) | 181 (13.6%) | <0.001 |
| Stridor at rest | 126 (1.0%) | 65 (0.6%) | 61 (4.7%) | <0.001 |
| Unable to feed | 187 (1.5%) | 74 (0.7%) | 113 (8.5%) | <0.001 |
| Lethargy or unconscious | 58 (0.5%) | 29 (0.3%) | 29 (2.2%) | <0.001 |
| Convulsions | 62 (0.5%) | 35 (0.3%) | 27 (2.0%) | <0.001 |
| Severe respiratory distress, n (%) | 2,352 (18.5%) | 1,667 (14.6%) | 685 (51.6%) | <0.001 |
| Grunting | 399 (3.1%) | 179 (1.6%) | 220 (16.7%) | <0.001 |
| Head nodding | 636 (5.0%) | 402 (3.5%) | 234 (17.8%) | <0.001 |
| Nasal flaring | 1,219 (9.6%) | 776 (6.8%) | 443 (33.5%) | <0.001 |
| Severe fast breathing<sup>1</sup> | 993 (7.8%) | 739 (6.5%) | 254 (19.4%) | <0.001 |
| Central cyanosis, n (%) | 104 (0.8%) | 26 (0.2%) | 78 (6.0%) | <0.001 |
| Temperature $\geq 38^{\circ}$ C, n (%) | 3,540 (28.4%) | 3,009 (26.8%) | 531 (41.9%) | <0.001 |
| Chest indrawing, n (%) | 3,944 (31.0%) | 3,126 (27.5%) | 818 (61.6%) | <0.001 |
| Wheezing, n (%) | 462 (3.6%) | 248 (2.2%) | 214 (16.1%) | <0.001 |
| Non-severe IMCI pneumonia<sup>2</sup>, n (%) | 11,514 (90.8%) | 10,449 (91.9%) | 1,065 (81.4%) | <0.001 |
| Severe IMCI pneumonia<sup>3</sup>, n (%) | 1,198 (9.4%) | 935 (8.2%) | 263 (19.8%) | <0.001 |

SpO<sub>2</sub> indicates peripheral arterial oxyhemoglobin saturation; IQR, interquartile range; WHO, World Health Organization; IMCI, Integrated Management of Childhood Illnesses.
<sup>1</sup>Respiratory rate ≥ 70 breaths per minute for 3-11 month olds, ≥ 60 breaths per minute for 12-59 month olds.
<sup>2</sup>Cough and/or difficult breathing plus fast breathing for age (respiratory rate ≥ 50 breaths per minute for 3-11 month olds, ≥ 40 breaths per minute for 12-59 month olds) or chest wall indrawing without any WHO danger signs (stridor at rest, unable to feed, lethargy or unconscious, convulsions).
<sup>3</sup>Cough and/or difficult breathing plus at least one WHO danger sign (stridor at rest, unable to feed, lethargy or unconscious, convulsions), WAZ <-3, or HIV-infection with chest indrawing.

We assessed the relationship between referral criteria and hypoxemia at SpO_2_<90%, 90%-92%, and <93% for the IMCI and WHO-composite models (Table 2). WAZ ≤-3, in both models, was associated with a SpO_2_ 90-92% and <93% but not <90%. In the WHO-composite model severe respiratory distress was associated with an increased adjusted odds of an abnormal SpO_2_ regardless of SpO_2_ range. While in the IMCI model danger signs were associated with hypoxemia, including respiratory distress in the WHO-composite model attenuated its effect at each SpO_2_ threshold. Respiratory distress was associated with each SpO_2_ threshold in the WHO-composite model.

**Table 2.**
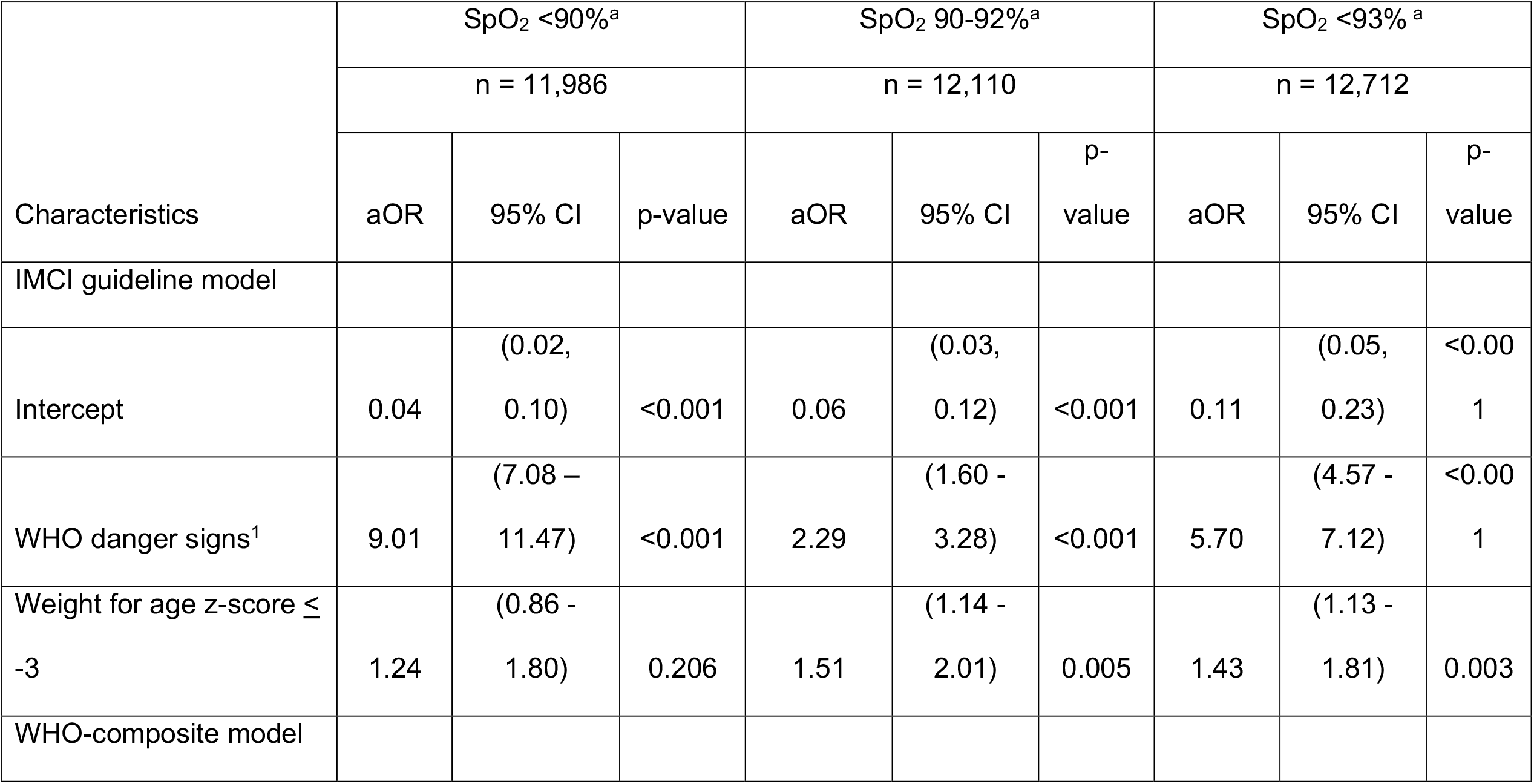
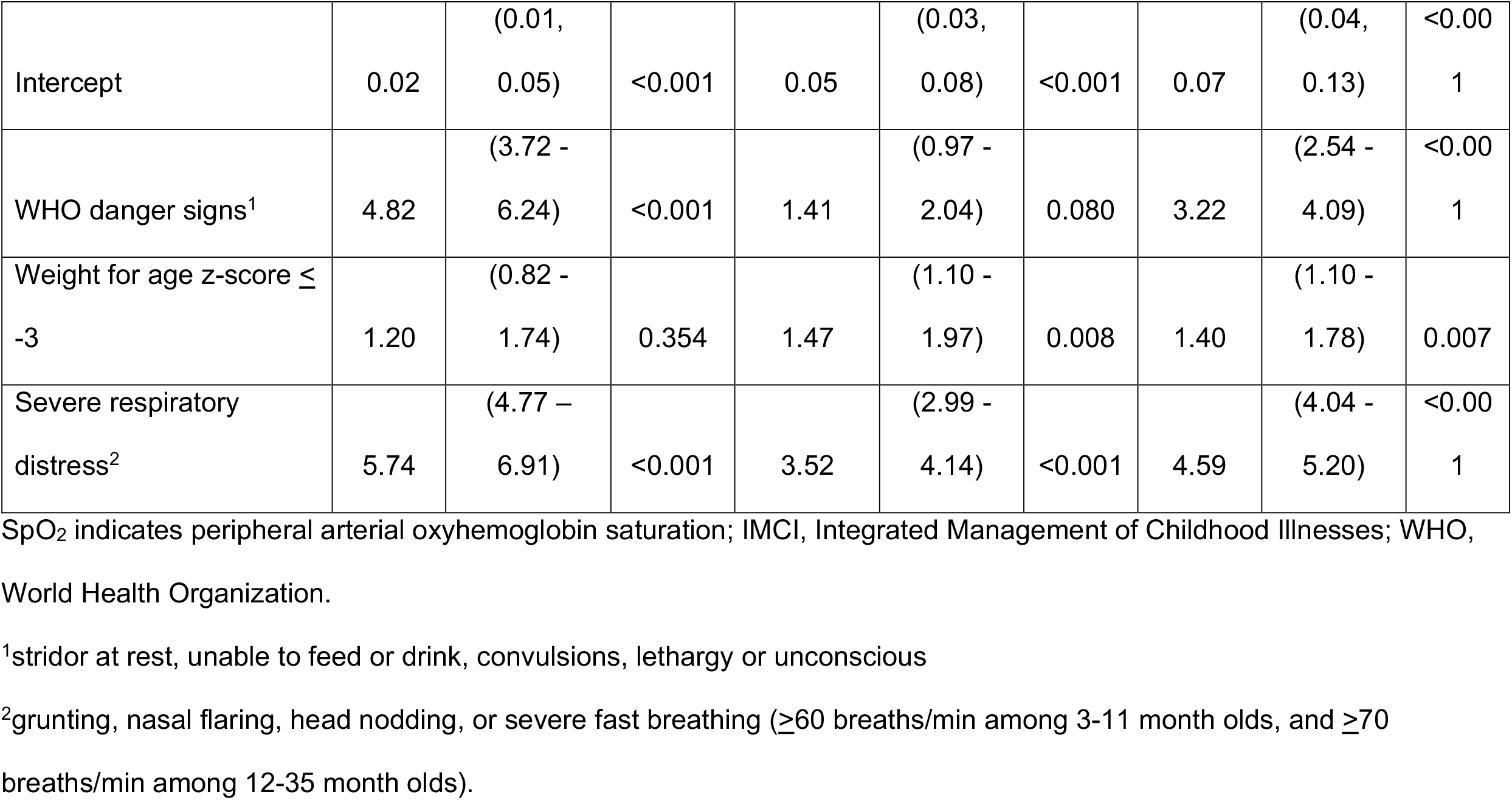
Association of IMCI guideline and WHO-composite models with SpO_2_ ranges (full dataset, N= 12,712)

### Multivariable predictive model comparison and score development

Derivation (n=3,813) and validation (n=8,899) datasets have similar patient characteristic distributions (Supplemental Material, all p>0.05). Table 3 presents adjusted odds ratios (aORs) for SpO_2_<93% using model-specific predictors from the derivation dataset. In the independent LASSO model, individual predictor scores ranged from -1 to 3. The composite LASSO model scores ranged from -1 to 4. Overall predictive performance of the independent (C-statistic=0.774) and composite LASSO models (C-statistic=0.773) was moderate (Table 3) and did not differ in discriminatory power (p=0.480), with total score ranges in the Supplemental Material. Model fit improved from the IMCI (BIC 5,536.5) and WHO-composite models (BIC 5,155.9) to the independent (BIC 4,650.6) and composite LASSO models (BIC 4,712.2). Overall, the independent (C-statistic=0.774) and composite LASSO models (C-statistic=0.773) better discriminated between children with and without a SpO_2_<93% than the IMCI (C-statistic=0.661) and WHO-composite models (C-statistic=0.734). SpO_2_<90% score development and cross-model comparison are in the Supplemental Material.

**Table 3.**
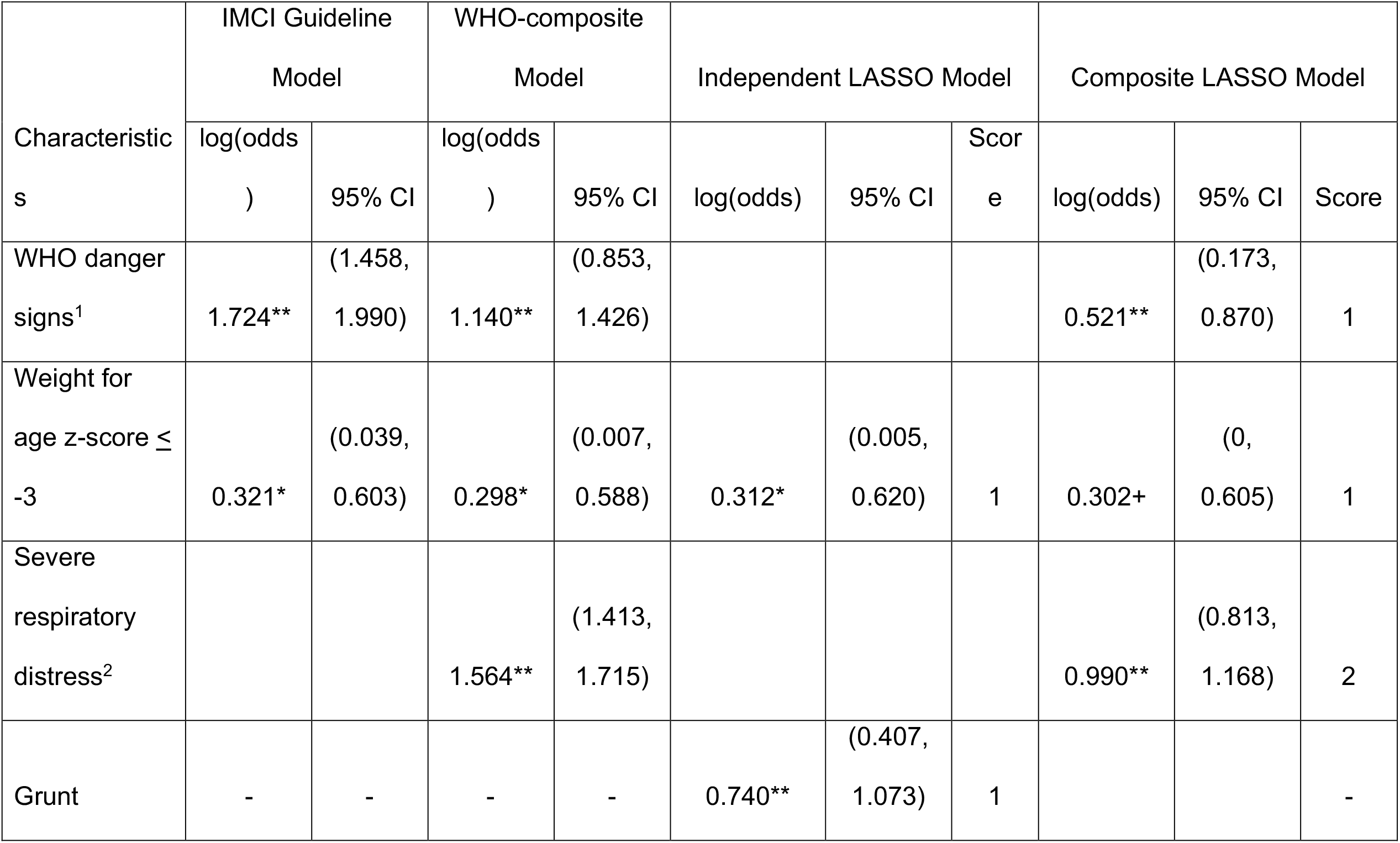
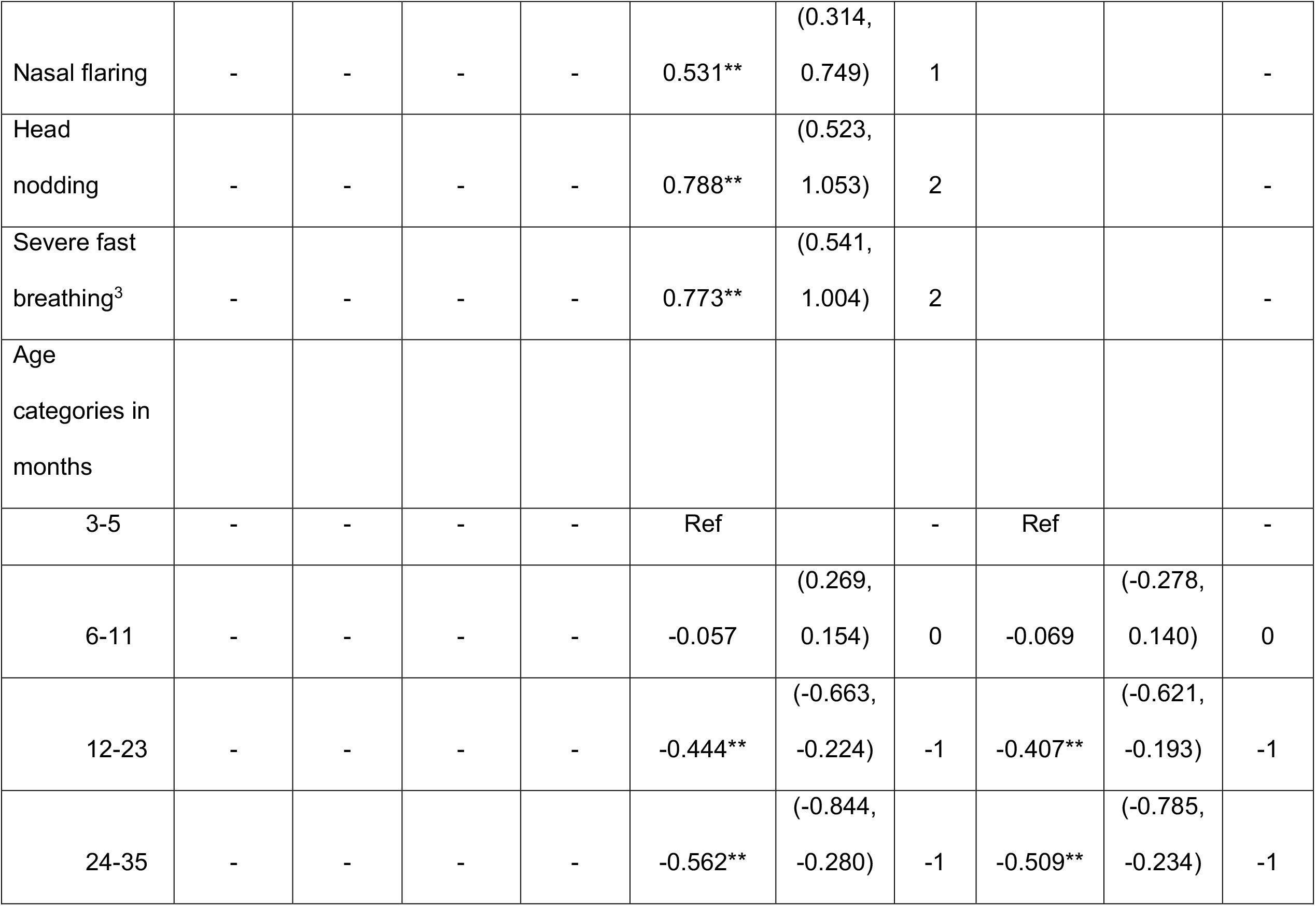
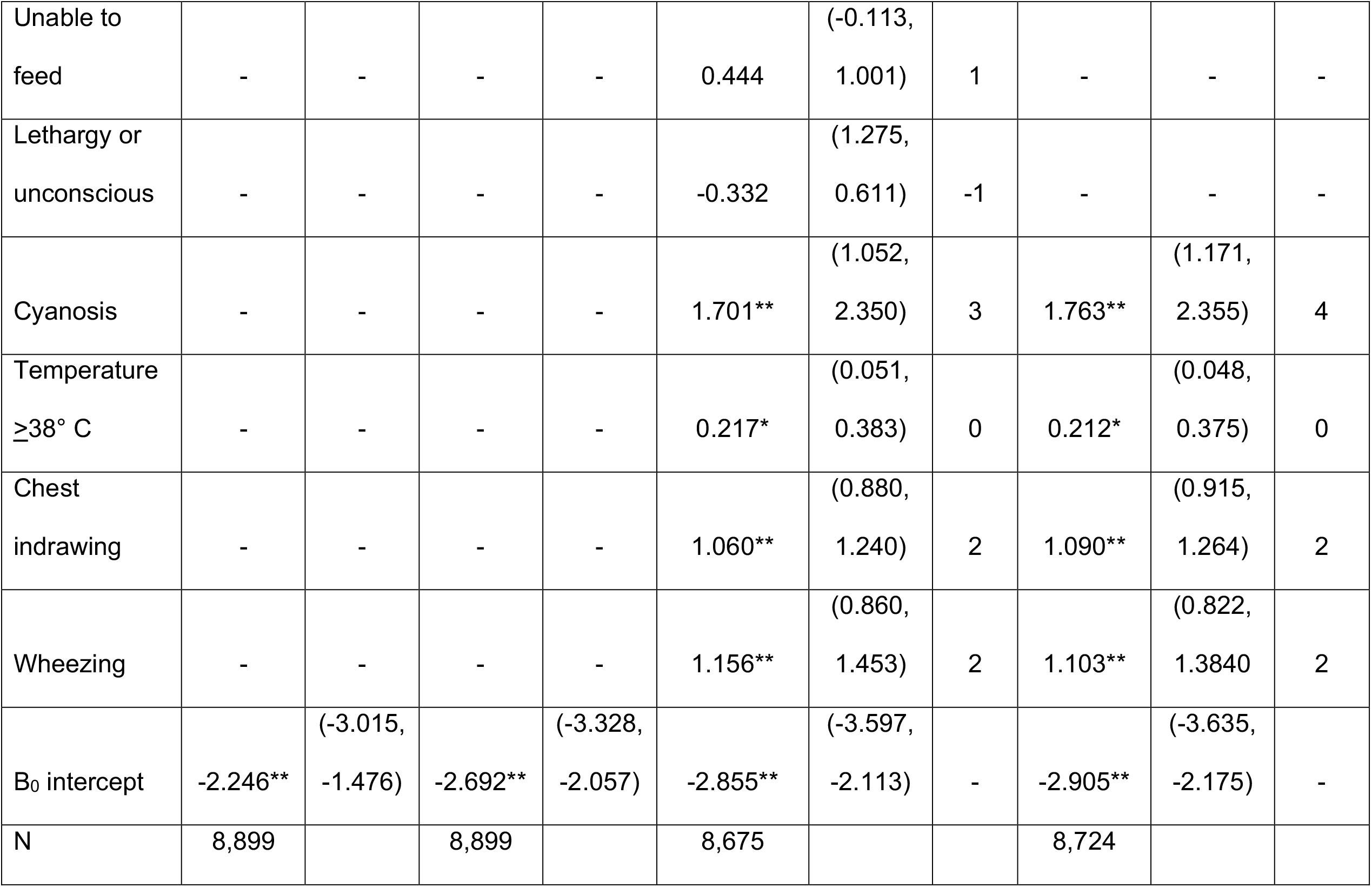
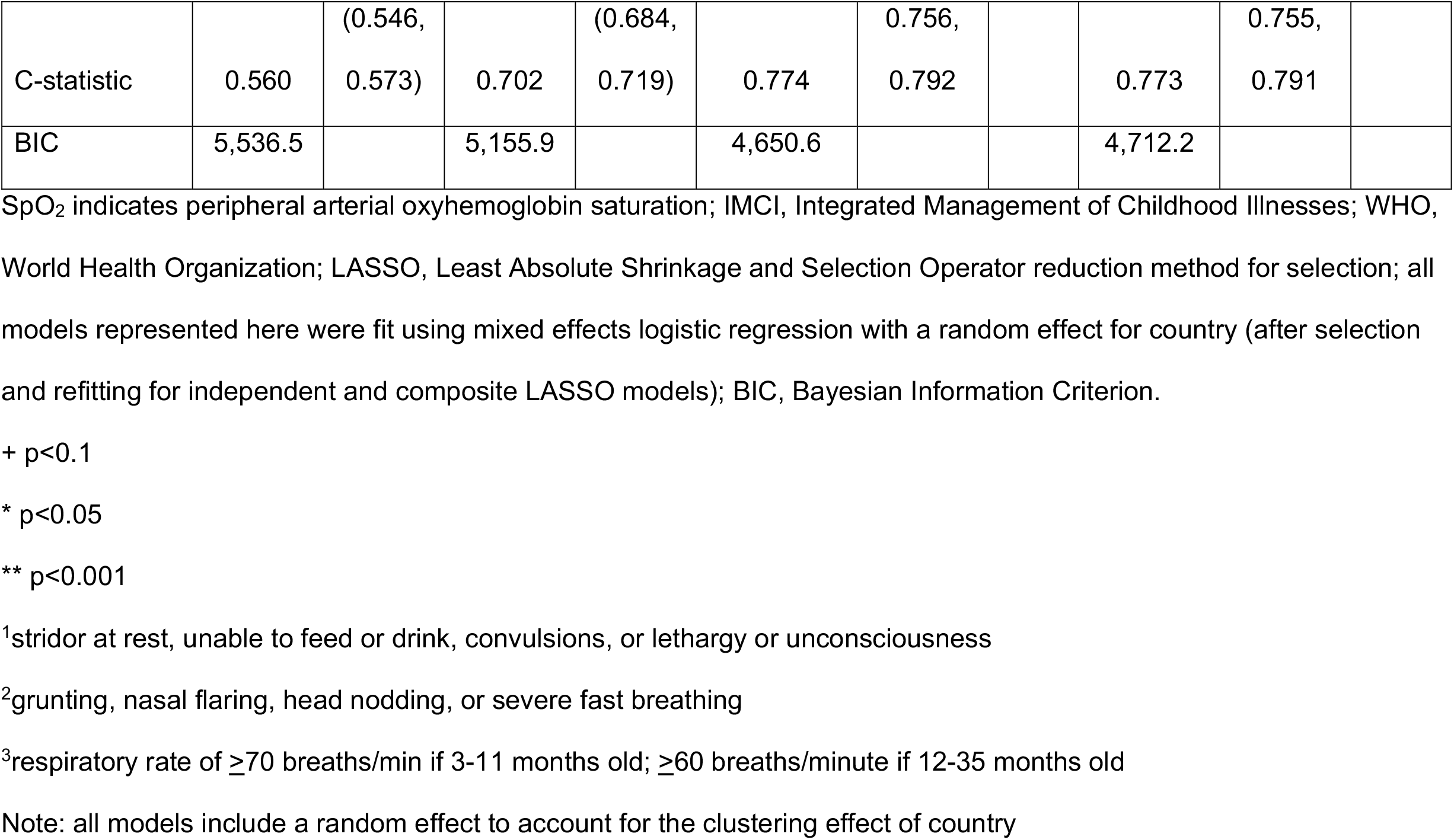
Clinical hypoxemia score for identifying a SpO_2_ <93% (derivation dataset)

### Clinical score performance – validation dataset

In the validation dataset, independent (C-statistic=0.745) and composite LASSO model (C-statistic=0.752) scores moderately discriminated between SpO_2_<93% and ≥94% cases (Figure 1). When examining the C-statistics adjusted for optimism, there was minimal C-statistic change for both scores (Table 4), and independent and composite LASSO model discriminatory power did not differ (p=0.480). For the independent and composite LASSO models about 4% of children with a score in the first stratum had SpO_2_<93% compared to >35% in the last stratum (Table 4).

**Table 4.**
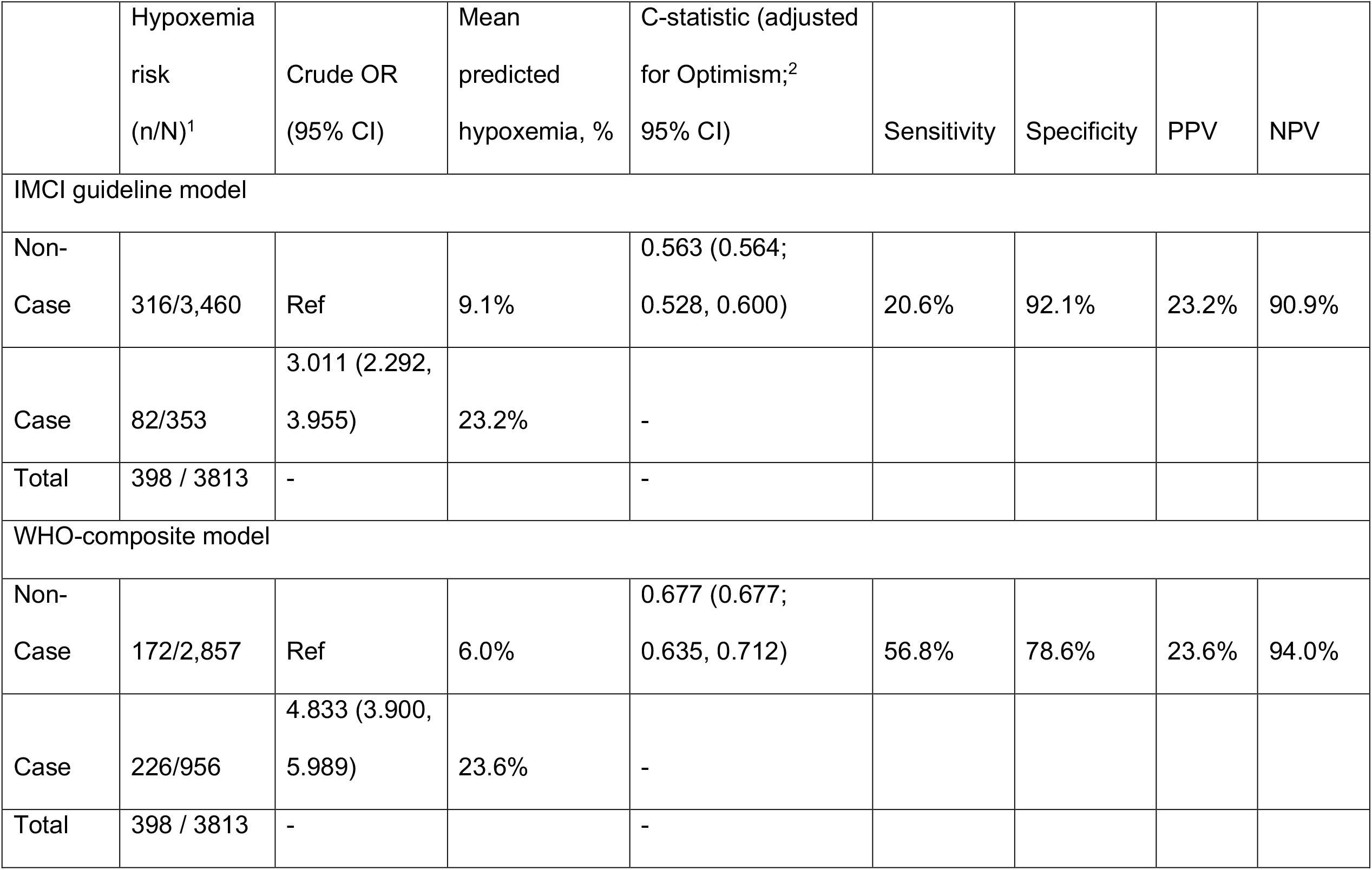
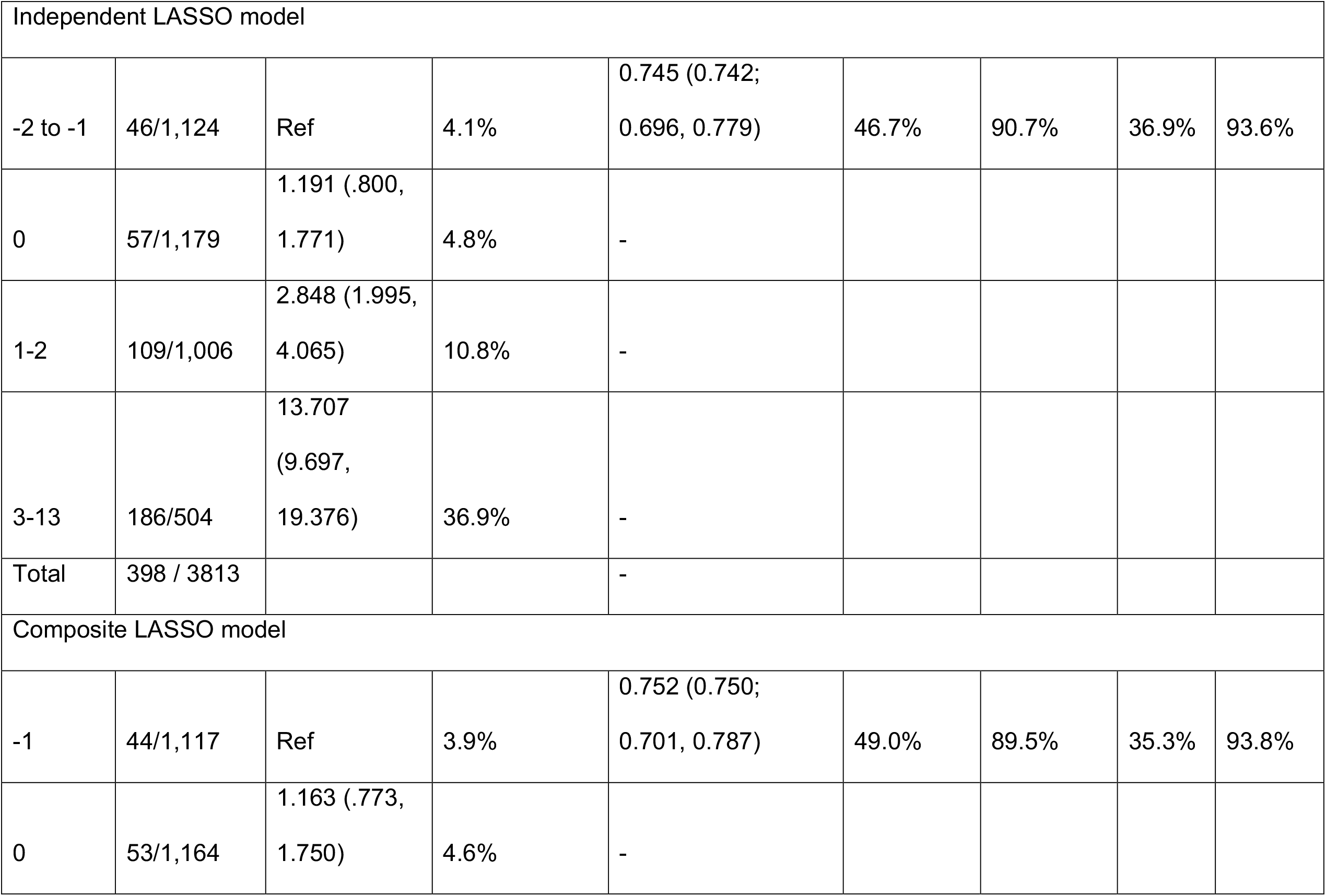
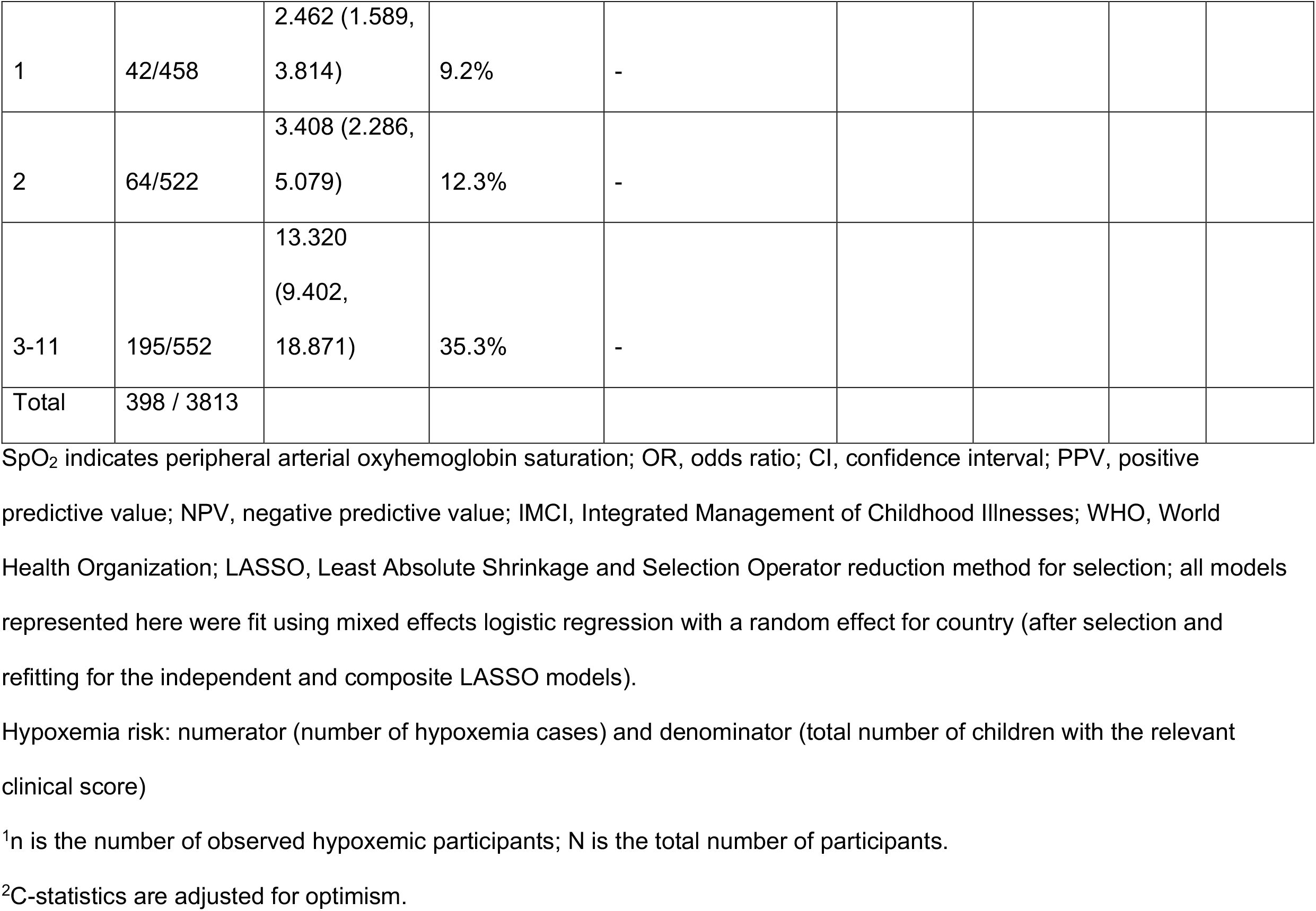
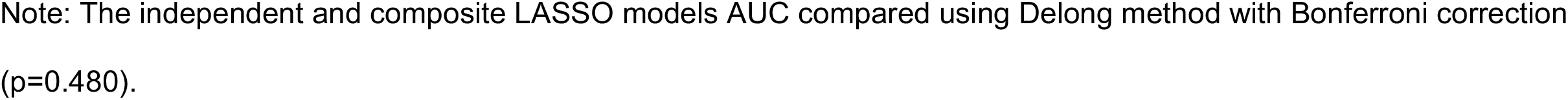
Model performance for identifying a SpO_2_<93% (validation dataset)

**Figure 1.**
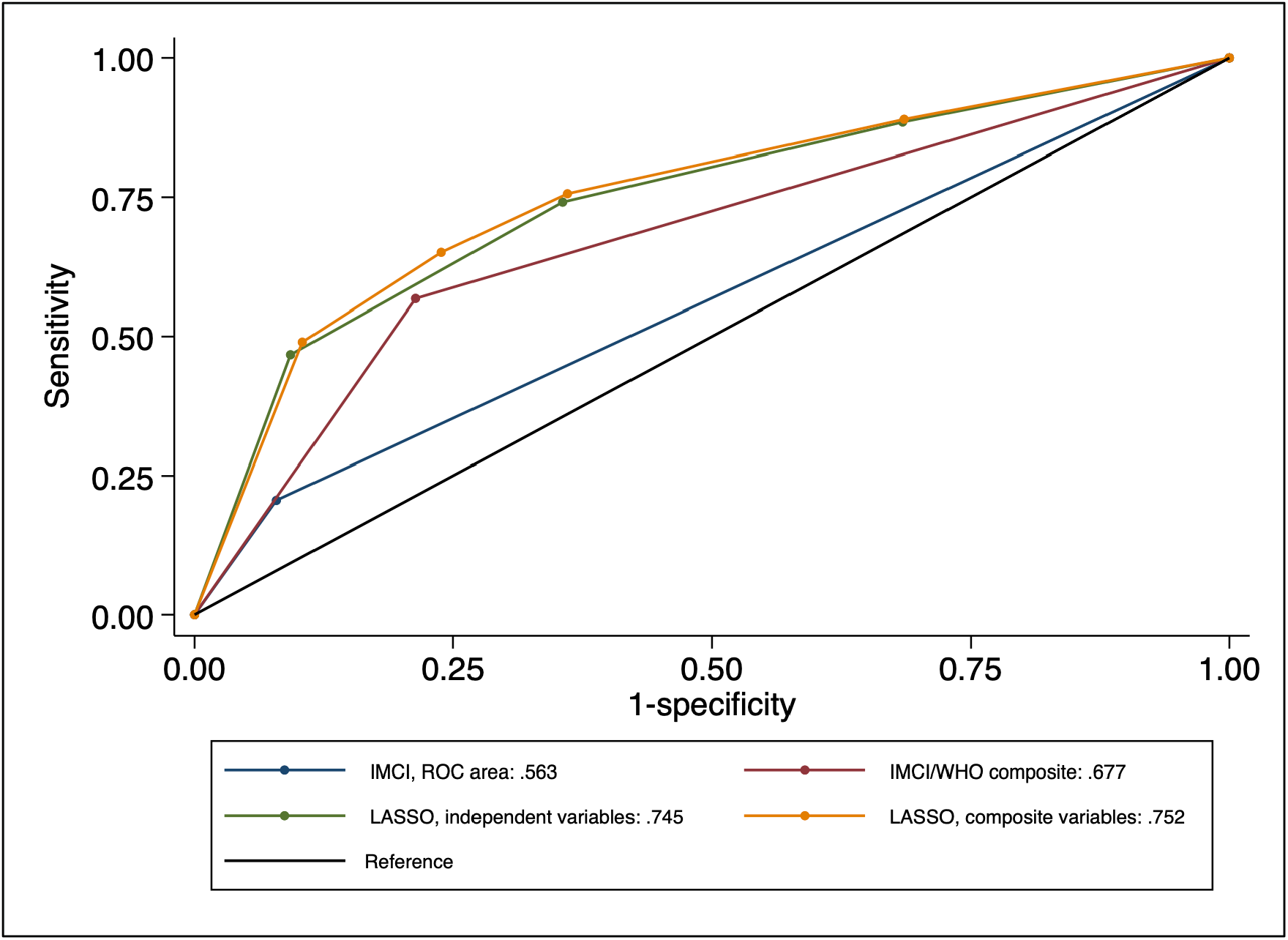
Comparison of ROC curves for identifying SpO_2_<93% cases (validation dataset) ROC indicates Receiver Operating Characteristic Curve; SpO_2_, peripheral arterial oxyhemoglobin saturation; IMCI, Integrated Management of Childhood Illnesses; WHO, World Health Organization; LASSO, Least Absolute Shrinkage and Selection Operator reduction method.

### Cross-model comparison: discriminating between hypoxemic and non-hypoxemic children

Among IMCI and WHO-composite model classified cases, 23% and 24% had a SpO_2_<93%, while among non-cases 9% and 6% had a SpO_2_<93% (Table 4). The ability to discriminate between SpO_2_<93% and ≤94% cases using the IMCI (C-statistic=0.563) and WHO-composite model (C-statistic=0.677) criteria was low. Based on model fit and discrimination, the composite LASSO model was the most predictive of SpO_2_<93%, identifying 49.0% of SpO_2_<93% cases. The independent LASSO model performed similarly, followed by WHO-composite and IMCI models. SpO_2_<90% results on score validation and ability to discriminate hypoxemia from non-hypoxemia cases are in the Supplemental Material.

### Clinical signs for a SpO_2_ <93%

The Supplemental Material includes the diagnostic performance of individual clinical signs for a SpO_2_<93% and <90% using the validation datasets.

## DISCUSSION

Using pooled data from 12,712 IMCI-defined child pneumonia cases evaluated at 21 clinics in Malawi and Bangladesh we examined WHO IMCI guideline hypoxemia identification performance and developed and internally validated clinical hypoxemia scores for use in LMICs where pulse oximeters suitable for pediatric outpatient care are scarce. Our findings suggest more hypoxemic children could be identified during outpatient care lacking pulse oximeters if additional signs of respiratory distress are incorporated into IMCI. Notably, >80% of hypoxemic cases with a SpO_2_<93% and >70% with a SpO_2_<90% were misclassified by IMCI as ineligible for hospital referral when pulse oximeters are unavailable. The independent LASSO model added age, severe respiratory distress, chest indrawing, cyanosis, fever, and wheezing parameters into the base IMCI guideline model and better identified children with a SpO_2_<93% than the IMCI and WHO-composite models. The composite LASSO model’s simplified composite variables may facilitate implementation. Both LASSO models also achieved excellent discrimination of SpO_2_<90% cases.

Unlike other studies evaluating hypoxemia predictors we focused on ambulatory rather than hospital settings. This distinction is important as our findings should therefore be generalizable to outpatient settings without oximeters and with lower hypoxemia prevalence than hospitals.^30,31^ Notably, individual variable sensitivity and specificity for hypoxemia are largely similar to hospital-based studies.^8^ However, our WHO-composite model and two LASSO models have good to excellent discriminatory value distinguishing between hypoxemic and non-hypoxemic pneumonia cases, contrasting to other work with smaller samples that limited analyses.^10^ An exception is a large, multi-center, hospital-based Nigerian study that found a combination of respiratory distress, inability to feed, cyanosis, lethargy and severe tachypnea had lower discrimination (C-statistic=0.655) for SpO_2_<90% amongst respiratory and non-respiratory cases than in our data.^9^ The lower C-statistic may reflect the authors inclusion of older children <15 years.

It is also important our results are interpreted within the broader child pneumonia context of known mortality risk factors. In the IMCI and WHO-composite models WAZ ≤-3 was not associated with a SpO_2_<90%, yet is a known mortality risk factor.^10,32^ Conversely wheezing was retained in both LASSO models but, when identified alone without accompanying respiratory distress, it is not associated with radiographic pneumonia or in-hospital mortality.^10,33^ Isolated wheeze usually reflects milder, self-limited viral illness (e.g., bronchiolitis) and is susceptible to non-differential misclassification when confused with transmitted upper respiratory sounds.

While our pooling of data from studies in South Asia and Africa aimed to improve generalizability, there are differences between the settings and studies that may be limit. Malawi data had a higher frequency of hypoxemia, danger signs, and respiratory distress than Bangladesh data. Alternatively, WAZ ≤-3 was more frequent in Bangladesh than Malawi. These differences, in part, may reflect higher HIV and malaria prevalence, modestly higher altitude in Malawi, or known challenges in identifying malnutrition in Malawi clinics.^34^ These diseases increase susceptibility for hypoxemia or, for malaria, increase the frequency of signs overlapping with IMCI pneumonia.^35^ Although the designs differed, all personnel were similarly trained per IMCI. The under 5 mortality rate in Malawi is higher than Bangladesh,^15,16^ which aligns with our results suggesting Malawi cases were more severe. Nevertheless, we attempted to account for unmeasured confounders from epidemiological, health system, and methodological differences by fitting models with a country variable.

In sum, these findings may improve hypoxemic pneumonia identification in clinics either without pulse oximetry at all or lacking pulse oximeters suitable for pediatric use. Given a SpO_2_ <93% is highly associated with mortality,^2,10,12^ earlier hypoxemic case identification with successful referral may reduce fatality. While these models could advance care for hypoxemic children they are an inadequate substitution for pulse oximeters. Pulse oximetry scale up in ambulatory settings must be prioritized, as should pulse oximeter device development targeted for young children in LMICs. Where pulse oximetry is unavailable for children, our models suggest children with chest indrawing and/or with other signs of respiratory distress could be referred. To successfully implement these models ambulatory health workers of varying pediatric experience will need further training to recognize signs of respiratory distress in children. Understanding how these changes may affect referral quality, uptake, and hospitalizations is critical. Next steps include external validation and research evaluating implementation feasibility of the hypoxemia scores.

## Data Availability

All data produced in the present study are available upon reasonable request to the authors

## Supplemental Material

Supplemental Figure 1: Study Definitions

Supplemental Figure 2: Study flowchart

Supplemental Figure 3. Comparison of ROC curves for identifying SpO_2_<90% cases (validation dataset)

Supplemental Table 1: Bangladesh: Risk factors for a SpO_2_<93%

Supplemental Table 2. Malawi: Risk factors for a SpO_2_<93%

Supplemental Table 3. Patient characteristics by SpO_2_<93%, stratified by development versus validation dataset

Supplemental Table 4. Model 3 (independent) scores and associated hypoxemia risk (SpO_2_<93%), sensitivity, specificity, and positive/negative likelihood ratios (validation dataset)

Supplemental Table 5. Model 4 (composite) scores and associated hypoxemia risk (SpO_2_<93%), sensitivity, specificity, and positive/negative likelihood ratios (validation dataset)

Supplemental Table 6. Association of Models with SpO_2_<90% and performance for identifying SpO_2_ <90% (development dataset)

Supplemental Table 7. Model 3 (independent) scores and associated hypoxemia risk (SpO_2_<90%), sensitivity, specificity, and positive/negative likelihood ratios (validation dataset)

Supplemental Table 8. Model 4 (composite) scores and associated hypoxemia risk (SpO_2_<90%), sensitivity, specificity, and positive/negative likelihood ratios (validation dataset)

Supplemental Table 9. Model performance and hypoxemia (SpO_2_<90%) case rate (validation dataset)

Supplemental Table 10. Performance of clinical signs for a SpO_2_<93% during outpatient pediatric care in Bangladesh and Malawi (validation dataset)

Supplemental Table 11. Performance of clinical signs for a SpO_2_<90% during outpatient pediatric care in Bangladesh and Malawi (validation dataset)

## Contributors

EDM had full access to data in the study and takes full responsibility for the integrity of the data and the accuracy of the data analysis. EDM conceived the study idea and HS, SH, and EDM designed the study; HS analyzed the data; SH, HS, and EDM drafted and revised the manuscript. All authors made substantial contributions to the interpretation of results, critical revision of the manuscript, and approved the final version of the manuscript.

## Acknowledgments

We offer our thanks to all of the caregivers and children participating in this research in Bangladesh and Malawi. In Bangladesh we would like to thank the Projahnmo Study Group field and data management staff, the Ministry of Health and Family Welfare, Government of Bangladesh. In Malawi we would like to thank Mr. Charles Makwenda and Rashid Deula for leading efforts in data collection and curation of the original study, the Republic of Malawi Ministry of Health and the communities and traditional authorities of Lilongwe and Mchinji districts of Malawi, and the dedication and hard work of our field and data collection staff. Lastly, we thank GlaxoSmithKline, Bill & Melinda Gates Foundation, and the National Institute of Health their support of this study.

