## Supplemental Material for "Clinical hypoxemia score for outpatient child pneumonia care lacking pulse oximetry in Africa and South Asia"

**Supplemental Material Contents:**

Supplemental Figure 1: Study Definitions – page 3

Supplemental Figure 2: Study flowchart – page 4

Supplemental Table 1: Bangladesh: Risk factors for a $SpO_2$<93% – page 5

Supplemental Table 2. Malawi: Risk factors for a $SpO_2$<93% – page 6

Supplemental Table 3. Patient characteristics by $SpO_2$<93%, stratified by development versus validation dataset – page 7

Supplemental Table 4. Independent LASSO model scores and associated hypoxemia risk ($SpO_2$<93%), sensitivity, specificity, and positive/negative likelihood ratios (validation dataset) – page 8

Supplemental Table 5. Composite LASSO model scores and associated hypoxemia risk ($SpO_2$<93%), sensitivity, specificity, and positive/negative likelihood ratios (validation dataset) – page 8

Supplemental Figure 3. Comparison of ROC curves for identifying $SpO_2$<90% cases (validation dataset) – page 9

Supplemental Table 6. Association of Models with $SpO_2$<90% and performance for identifying $SpO_2$ <90% (development dataset) – page 10

Supplemental Table 7. Independent LASSO model scores and associated hypoxemia risk ($SpO_2$<90%), sensitivity, specificity, and positive/negative likelihood ratios (validation dataset) – page 11

Supplemental Table 8. Composite LASSO model scores and associated hypoxemia risk ($SpO_2$<90%), sensitivity, specificity, and positive/negative likelihood ratios (validation dataset) – page 11

Supplemental Table 9. Model performance and hypoxemia ($SpO_2$<90%) case rate (validation dataset) – page 12

Supplemental Table 10. Performance of clinical signs for a $SpO_2$<93% during outpatient pediatric care in Bangladesh and Malawi (validation dataset) – page 13

Supplemental Table 11. Performance of clinical signs for a $SpO_2$<90% during outpatient pediatric care in Bangladesh and Malawi (validation dataset) – page 16

**Supplemental Figure 1. Study Definitions**

| | |
|---|---|
| Valid $SpO_2$ | Oxygen saturation measurement obtained within 5 minutes of measurement, with a stable and consistent plethysmography wave or signal IQ bar with two or more bars, and a stable oxygen saturation reading for at least three full seconds. |
| IMCI non-severe pneumonia | Cough and/or difficult breathing with fast breathing for age (breaths/min ≥50 for 3-11 months; ≥40 for 12-35 months) and/or chest indrawing |
| IMCI severe pneumonia | Cough and/or difficult breathing with WHO danger sign |
| Severe respiratory distress | grunting, nasal flaring, head nodding, or severe fast breathing |
| Severe fast breathing for age | breaths/min ≥70 for 3-11 months; ≥60 for 12-35 months |
| Fever | ≥38° C |
| WHO danger signs | stridor, unable to feed/drink, convulsions, or unconscious or lethargy |
| IMCI model | WHO-defined danger signs and weight for age z-score < -3 |
| WHO-composite model | WHO-defined danger signs, weight for age z-score < -3, severe respiratory distress |
| [*]Independent LASSO model | Weight for age z-score < -3, grunting, nasal flaring, head nodding, severe fast breathing, child age, cyanosis, inability to feed, lethargy or unconscious, presence of fever, chest indrawing, and wheezing |
| [*]Composite LASSO model | WHO danger signs, weight for age z-score < -3, severe respiratory distress, child age, cyanosis, fever, chest indrawing, wheezing |

*LASSO selection (Independent and Composite LASSO models): starting list included WHO danger signs (independent or composite), severe respiratory distress (independent or composite), central cyanosis, fever, chest indrawing, and wheezing. The reduction method selected the variables listed in Supplemental Figure 1. Reference to "WHO danger signs" or "severe respiratory distress" means use of the composite term/variable.

**Supplemental Figure 2. Study flowchart**

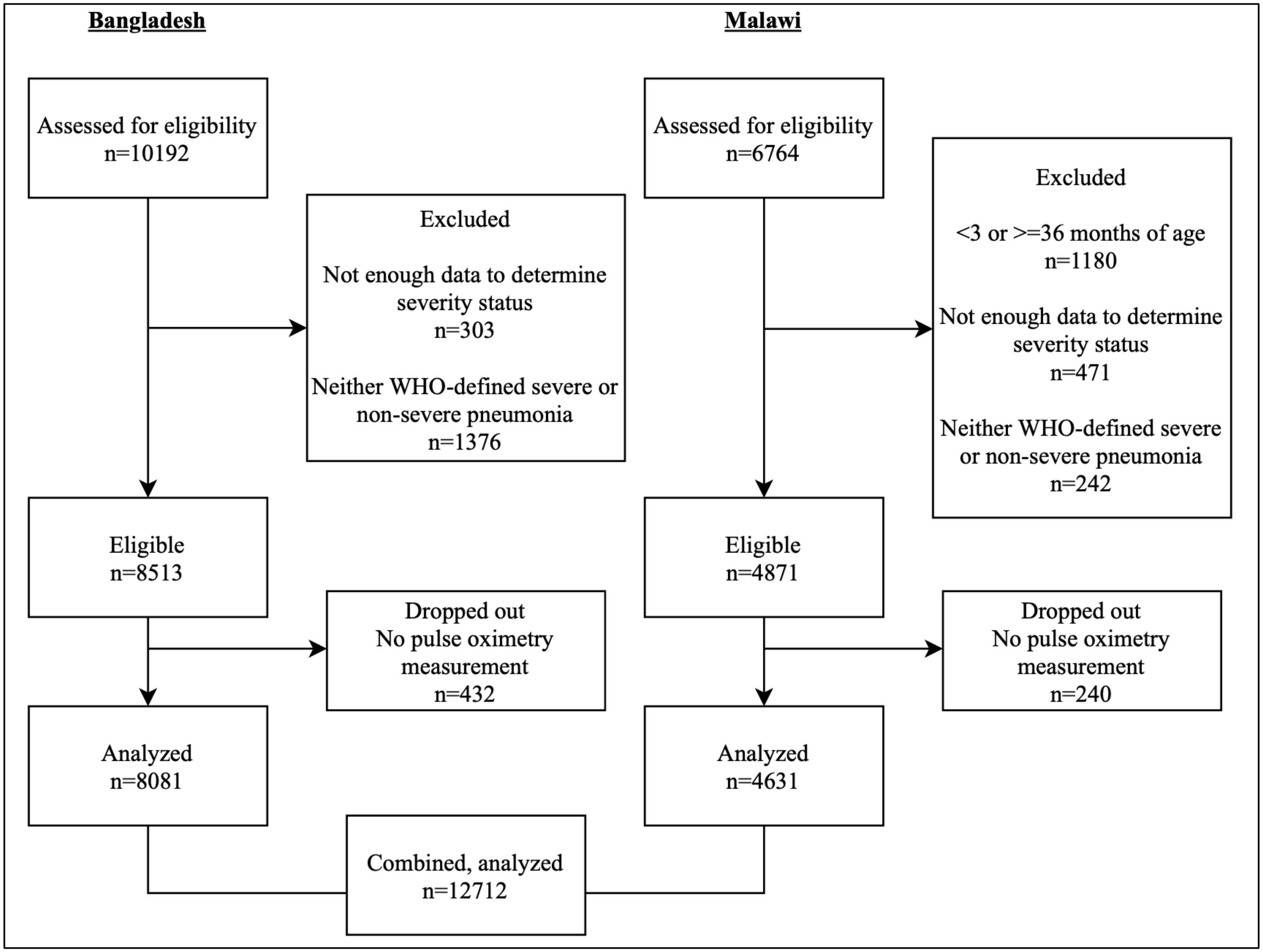

**Supplemental Table 1. Bangladesh: Risk factors for a $SpO_2$ <93% (full dataset, N=8,081)**

| | $SpO_2 \geq 93\%$ N=7,598 | $SpO_2$ <93% N=483 | Crude odds Ratio | 95% CI | p-value |
|---|---|---|---|---|---|
| Age (months) | | | | | |
| 3-5 | 1,617 (91.8%) | 144 (8.2%) | Ref | | |
| 6-11 | 2,249 (93.2%) | 164 (6.8%) | 0.82* | (0.65 - 1.03) | 0.092 |
| 12-23 | 2,500 (95.2%) | 127 (4.8%) | 0.57*** | (0.45 - 0.73) | <0.001 |
| 24-35 | 1,232 (96.2%) | 48 (3.8%) | 0.44*** | (0.31 - 0.61) | <0.001 |
| Child sex | | | | | |
| Male | 4,414 (94.4%) | 263 (5.6%) | Ref | | |
| Female | 3,184 (93.5%) | 220 (6.5%) | 1.16 | (0.96 - 1.40) | 0.116 |
| Weight for age z-score (WAZ) | | | | | |
| WAZ ≥ -2.0 | 5,387 (94.4%) | 322 (5.6%) | Ref | | |
| -3.0 < WAZ < -2.0 | 1,561 (94.1%) | 97 (5.9%) | 1.04 | (0.82 - 1.31) | 0.745 |
| WAZ ≤ -3.0 | 650 (91.0%) | 64 (9.0%) | 1.65*** | (1.24 - 2.18) | <0.001 |
| Fever | 1,907 (25.1%) | 196 (40.6%) | 2.04*** | (1.69 - 2.46) | <0.001 |
| WHO danger signs | 21 (0.3%) | 3 (0.6%) | 2.26 | (0.67 - 7.59) | 0.189 |
| Stridor | 0 (0.0%) | 0 (0.0%) | - | | - |
| Unable to feed | 0 (0.0%) | 0 (0.0%) | - | | - |
| Convulsions | 3 (0.0%) | 1 (0.2%) | 2.63 | (0.77 - 8.97) | 0.122 |
| Lethargy or unconscious | 18 (0.2%) | 3 (0.6%) | 5.25 | (0.55 - 50.59) | 0.151 |
| Severe respiratory distress | 851 (11.2%) | 186 (38.5%) | 5.24*** | (4.26 - 6.44) | <0.001 |
| Grunting | 26 (0.3%) | 13 (2.7%) | 8.06*** | (4.11 - 15.78) | <0.001 |
| Nasal flaring | 367 (4.8%) | 92 (19.0%) | 4.64*** | (3.61 - 5.95) | <0.001 |
| Head nodding | 317 (4.2%) | 96 (19.9%) | 5.70*** | (4.44 - 7.32) | <0.001 |
| Severe fast breathing[a] | 344 (4.5%) | 79 (16.4%) | 4.12*** | (3.17 - 5.37) | <0.001 |
| Central cyanosis | 7 (0.1%) | 7 (1.4%) | 15.95*** | (5.57 - 45.65) | <0.001 |
| Chest indrawing | 2,493 (32.8%) | 307 (63.6%) | 3.57*** | (2.95 - 4.33) | <0.001 |
| Wheezing | 25 (0.3%) | 12 (2.5%) | 7.72*** | (3.85 - 15.46) | <0.001 |

*** p<0.001, ** p<0.05, * p<0.1; Severe fast breathing defined as: ≥70 breaths/min among 3-11 month olds, and ≥60 breaths/min among 12-35 month olds; Fever defined as body temperature ≥38 degrees Celsius

**Supplemental Table 2. Malawi: Risk factors for a $SpO_2$ <93% (full dataset, N=4,631)**

| | **Not hypoxemic** N=3,786 | **Hypoxemic** N=845 | **Crude odds Ratio** | **95% CI** | **p-value** |
|---|---|---|---|---|---|
| Age (months) | | | | | |
| 3-5 | 617 (79.3%) | 161 (20.7%) | Ref | | |
| 6-11 | 1,108 (79.7%) | 283 (20.3%) | 0.98 | (0.79 - 1.22) | 0.847 |
| 12-23 | 1,404 (82.7%) | 294 (17.3%) | 0.80** | (0.65 - 0.99) | 0.044 |
| 24-35 | 657 (86.0%) | 107 (14.0%) | 0.62*** | (0.48 - 0.82) | 0.001 |
| Child sex | | | | | |
| Male | 1,793 (80.8%) | 425 (19.2%) | Ref | | |
| Female | 1,766 (83.1%) | 360 (16.9%) | 0.86* | (0.74 - 1.00) | 0.057 |
| Missing | 227 (79.1%) | 60 (20.9%) | | | |
| Weight for age z-score (WAZ) | | | | | |
| WAZ ≥ -2.0 | 3,076 (82.4%) | 656 (17.6%) | Ref | | |
| -3.0 < WAZ < -2.0 | 338 (80.1%) | 84 (19.9%) | 1.17 | (0.90 - 1.50) | 0.237 |
| WAZ ≤ -3.0 | 110 (79.7%) | 28 (20.3%) | 1.19 | (0.78 - 1.82) | 0.413 |
| Missing | 262 (77.3%) | 77 (22.7%) | | | |
| Fever, temp>=38c | 1,102 (30.5%) | 335 (42.8%) | 1.71*** | (1.46 - 2.00) | <0.0001 |
| WHO danger signs | 162 (4.3%) | 178 (21.1%) | 5.97*** | (4.75 - 7.50) | <0.0001 |
| Stridor | 65 (1.7%) | 61 (7.5%) | 4.56*** | (3.19 - 6.53) | <0.0001 |
| Unable to feed | 74 (2.0%) | 113 (13.5%) | 7.78*** | (5.75 - 10.55) | <0.0001 |
| Convulsions | 17 (0.5%) | 24 (2.9%) | 6.54*** | (3.50 - 12.22) | <0.0001 |
| No movement / lethargy | 26 (0.7%) | 28 (3.3%) | 4.99*** | (2.91 - 8.55) | <0.0001 |
| Severe respiratory distress | 816 (21.6%) | 499 (59.1%) | 7.14*** | (6.06 - 8.41) | <0.0001 |
| Grunting | 153 (4.1%) | 207 (24.9%) | 7.83*** | (6.25 - 9.82) | <0.0001 |
| Nasal flaring | 409 (10.8%) | 351 (41.9%) | 5.93*** | (5.00 - 7.04) | <0.0001 |
| Head nodding | 85 (2.3%) | 138 (16.6%) | 8.62*** | (6.50 - 11.43) | <0.0001 |
| Severe fast breathing | 395 (10.5%) | 175 (21.2%) | 2.30*** | (1.89 - 2.80) | <0.0001 |
| Central cyanosis | 19 (0.5%) | 71 (8.6%) | 18.49*** | (11.08 - 30.85) | <0.0001 |
| Chest indrawing | 633 (16.7%) | 511 (60.5%) | 7.64*** | (6.49 - 8.98) | <0.0001 |
| Wheezing | 223 (5.9%) | 202 (23.9%) | 5.02*** | (4.08 - 6.18) | <0.0001 |

*** p<0.001, ** p<0.05, * p<0.1; Severe fast breathing defined as: ≥70 breaths/min among 3-11 month olds, and ≥60 breaths/min among 12-35 month olds; Fever defined as body temperature ≥38 degrees Celsius

**Supplemental Table 3. Patient characteristics by $SpO_2$<93%, stratified by development versus validation dataset**

| | Total | Development | Validation | p-value |
|---|---|---|---|---|
| | N=12,712 | N=8,899 | N=3,813 | |
| Study country | | | | 1.00 |
| Bangladesh | 8,081 (63.6%) | 5,657 (63.6%) | 2,424 (63.6%) | |
| Malawi | 4,631 (36.4%) | 3,242 (36.4%) | 1,389 (36.4%) | |
| Child age (months) | 12 (6-19) | 11 (6-19) | 12 (6-19) | 0.20 |
| Child sex | | | | 0.41 |
| Male | 6,895 (54.2%) | 4,802 (54.0%) | 2,093 (54.9%) | |
| Female | 5,530 (43.5%) | 3,890 (43.7%) | 1,640 (43.0%) | |
| Missing | 287 ( 2.3%) | 207 ( 2.3%) | 80 ( 2.1%) | |
| Weight for age z-score (WAZ) | | | | 0.66 |
| WAZ ≥ -2.0 | 9,441 (74.3%) | 6,585 (74.0%) | 2,856 (74.9%) | |
| -3.0 < WAZ < -2.0 | 2,080 (16.4%) | 1,468 (16.5%) | 612 (16.1%) | |
| WAZ ≤ -3.0 | 852 ( 6.7%) | 603 ( 6.8%) | 249 ( 6.5%) | |
| Missing | 339 ( 2.7%) | 243 ( 2.7%) | 96 ( 2.5%) | |
| WHO danger signs | 364 ( 2.9%) | 252 ( 2.8%) | 112 ( 2.9%) | 0.74 |
| Stridor | 126 ( 1.0%) | 84 ( 0.9%) | 42 ( 1.1%) | 0.41 |
| Unable to feed | 187 ( 1.5%) | 124 ( 1.4%) | 63 ( 1.7%) | 0.27 |
| Lethargy | 58 ( 0.5%) | 41 ( 0.5%) | 17 ( 0.4%) | 0.91 |
| Seizures | 62 ( 0.5%) | 46 ( 0.5%) | 16 ( 0.4%) | 0.47 |
| Severe respiratory distress | 2,352 (18.5%) | 1,630 (18.3%) | 722 (18.9%) | 0.41 |
| Grunting | 399 ( 3.1%) | 273 ( 3.1%) | 126 ( 3.3%) | 0.49 |
| Head nodding | 636 ( 5.0%) | 440 ( 5.0%) | 196 ( 5.2%) | 0.65 |
| Nasal flaring | 1,219 ( 9.6%) | 855 ( 9.6%) | 364 ( 9.6%) | 0.91 |
| Severe fast breathing | 993 ( 7.8%) | 687 ( 7.7%) | 306 ( 8.1%) | 0.55 |
| Central cyanosis | 104 ( 0.8%) | 73 ( 0.8%) | 31 ( 0.8%) | 0.96 |
| Fever | 3,540 (28.4%) | 2,477 (28.3%) | 1,063 (28.5%) | 0.81 |
| Chest indrawing | 3,944 (31.0%) | 2,751 (30.9%) | 1,193 (31.3%) | 0.68 |
| Wheezing | 462 ( 3.6%) | 319 ( 3.6%) | 143 ( 3.8%) | 0.65 |
| Non-severe pneumonia | 11,514 (90.8%) | 8,054 (90.7%) | 3,460 (91.1%) | 0.55 |
| Severe pneumonia | 1,198 ( 9.4%) | 845 ( 9.5%) | 353 ( 9.3%) | 0.67 |

Severe fast breathing defined as: ≥70 breaths/min among 3-11 month olds, and ≥60 breaths/min among 12-35 month olds; Fever defined as body temperature ≥38 degrees Celsius

**Supplemental Table 4. Independent LASSO model scores and associated hypoxemia risk ($SpO_2$<93%), sensitivity, specificity, and positive/negative likelihood ratios (validation dataset)**

| Score | Hypoxemia risk | Sensitivity | Specificity | LR+ | LR- |
|---|---|---|---|---|---|
| -2 | 0 | 0 | 100 | 0.00 | 1.00 |
| -1 | 29 | 12 | 68 | 0.37 | 1.29 |
| 0 | 31 | 14 | 67 | 0.44 | 1.28 |
| 1 | 12 | 11 | 87 | 0.85 | 1.02 |
| 2 | 14 | 17 | 86 | 1.22 | 0.97 |
| 3 | 4 | 8 | 96 | 2.05 | 0.96 |
| 4 | 4 | 17 | 97 | 5.78 | 0.86 |
| 5 | 2 | 9 | 99 | 6.67 | 0.92 |
| 6 | 1 | 5 | 99 | 9.53 | 0.95 |
| 7 | 1 | 4 | 100 | 10.73 | 0.97 |
| 8 | 0 | 2 | 100 | 17.16 | 0.98 |
| 9 | 0 | 1 | 100 | 17.16 | 0.99 |
| 10 | 0 | 1 | 100 | . | 0.99 |
| 11 | 0 | 0 | 100 | 8.58 | 1.00 |

**Supplemental Table 5. Composite LASSO model scores and associated hypoxemia risk ($SpO_2$<93%), sensitivity, specificity, and positive/negative likelihood ratios (validation dataset)**

| Score | Hypoxemia risk | Sensitivity | Specificity | LR+ | LR- |
|---|---|---|---|---|---|
| -1 | 29 | 11 | 69 | 0.35 | 1.30 |
| 0 | 31 | 13 | 67 | 0.41 | 1.28 |
| 1 | 12 | 11 | 88 | 0.87 | 1.02 |
| 2 | 14 | 16 | 87 | 1.20 | 0.97 |
| 3 | 5 | 14 | 96 | 3.13 | 0.90 |
| 4 | 6 | 16 | 95 | 3.37 | 0.89 |
| 5 | 2 | 9 | 99 | 10.80 | 0.92 |
| 6 | 1 | 6 | 100 | 12.58 | 0.95 |
| 7 | 0 | 2 | 100 | 13.73 | 0.98 |
| 8 | 0 | 2 | 100 | 25.74 | 0.99 |
| 9 | 0 | 2 | 100 | 60.06 | 0.98 |
| 10 | 0 | 1 | 100 | 17.16 | 1.00 |

Supplemental Figure 3. Comparison of ROC curves for identifying $SpO_2$ <90% cases (validation dataset)

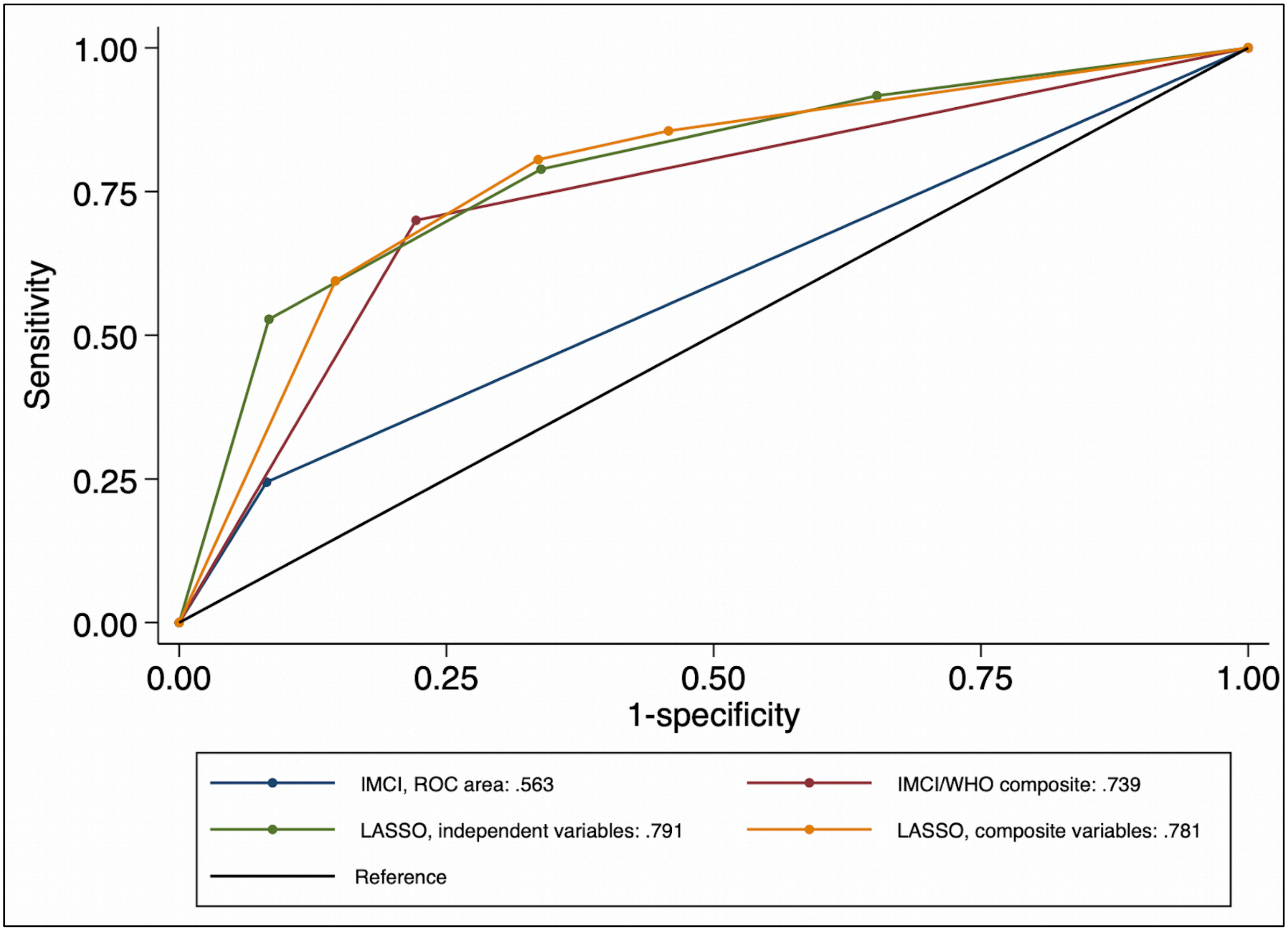

**Supplemental Table 6. Association of Models with $SpO_2$ <90% and performance for identifying $SpO_2$ <90% (development dataset)**

| | IMCI model | | WHO-composite model | | Independent LASSO model | | | Composite LASSO model | | |
|---|---|---|---|---|---|---|---|---|---|---|
| Predictors | log(odds) | 95% CI | log(odds) | 95% CI | log(odds) | 95% CI | Score | log(odds) | 95% CI | Score |
| WHO danger sign, yes | 2.247** | (1.964,2.530) | 1.649** | (1.345,1.953) | - | - | - | 1.005** | (.626,1.384) | 2 |
| WAZ < -3, yes | 0.290 | (-.140,.720) | 0.297 | (-0.139,.733) | - | - | - | - | - | - |
| Severe respiratory distress | - | - | 1.711** | (1.490,1.931) | - | - | - | 1.054** | (.789,1.319) | 2 |
| Grunt | - | - | - | - | 0.581** | (.204,.959) | 1 | | | - |
| Nasal flaring | - | - | - | - | 0.599** | (.309,.890) | 1 | | | - |
| Head nodding | - | - | - | - | 0.949** | (.621,1.278) | 2 | | | - |
| Severe fast breathing | - | - | - | - | 0.907** | (.600,1.215) | 2 | | | - |
| Age (months) | - | - | - | - | - | - | - | | | - |
| 3-5 | - | - | - | - | Ref | | - | Ref | | - |
| 6-11 | - | - | - | - | -0.111 | (-.420,.198) | 0 | -1.02 | (-.403,.199) | -2 |
| 12-23 | - | - | - | - | -0.608** | (-.936,-.280) | -1 | -0.532** | (-.847,-.217) | -1 |
| 24-35 | - | - | - | - | -0.738** | (-1.168,-.308) | -1 | -0.587** | (-.996,-.177) | -1 |
| Unable to feed | - | - | - | - | 0.351 | (-0.210,.911) | 1 | - | - | - |
| No movement / lethargy | - | - | - | - | -0.332 | (-1.275,0.611) | -1 | - | - | - |
| Cyanosis | - | - | - | - | 2.503** | (1.874,3.133) | 5 | 2.224** | (1.645,2.802) | 4 |
| Fever | - | - | - | - | 0.186 | (-.060,.432) | 0 | 0.220+ | (-.019,.459) | 0 |
| Chest indrawing | - | - | - | - | 1.459** | (1.171,1.747) | 3 | 1.467** | (1.191,1.743) | 3 |
| Wheezing | - | - | - | - | - | - | - | - | - | - |
| $B_0$, intercept | -3.272** | (-4.185,-2.360) | -3.838** | (-4.580,-3.097) | -4.110 | (-5.081,-3.138) | - | -4.168** | (-5.082,-3.254) | - |
| N | 8899 | | 8899 | | 8669 | | | 8713 | | |
| C-statistic | 0.612 | (0.589, 0.635) | 0.761 | (0.736, 0.786) | 0.832 | (0.809, 0.855) | | 0.834 | (0.812, 0.856) | |
| BIC | 2995.3 | | 2776.5 | | 2350.1 | | | 2423.9 | | |

+ $p<0.1$, * $p<0.05$, ** $p<0.001$ Severe respiratory distress: grunting, nasal flaring, head nodding, or severe fast breathing; WHO danger signs: stridor, unable to feed/drink, convulsions, or lethargy or unconsciousness. Severe fast breathing: breaths/min ≥70 3-11months; ≥60 12-35months. Fever: ≥38. "-" indicates where variable is not specified. LASSO, Least Absolute Shrinkage and Selection Operator reduction method for selection; all models represented here were fit using mixed effects logistic regression with a random effect for country (after selection and refitting for Model 3 and 4).

**Supplemental Table 7. Independent LASSO model scores and associated hypoxemia risk ($SpO_2$<90%), sensitivity, specificity, and positive/negative likelihood ratios (validation dataset)**

| Score | Hypoxemia risk | Sensitivity | Specificity | LR+ | LR- |
|---|---|---|---|---|---|
| -2 | 0 | 0 | 100 | 0.00 | 1.00 |
| -1 | 33 | 8 | 65 | 0.24 | 1.40 |
| 0 | 31 | 13 | 69 | 0.41 | 1.27 |
| 1 | 4 | 6 | 96 | 1.36 | 0.98 |
| 2 | 9 | 8 | 91 | 0.86 | 1.01 |
| 3 | 12 | 13 | 88 | 1.03 | 1.00 |
| 4 | 4 | 9 | 96 | 2.36 | 0.95 |
| 5 | 3 | 11 | 97 | 3.88 | 0.92 |
| 6 | 1 | 11 | 99 | 11.98 | 0.90 |
| 7 | 1 | 8 | 100 | 17.81 | 0.92 |
| 8 | 1 | 6 | 100 | 20.18 | 0.95 |
| 9 | 0 | 2 | 100 | 80.73 | 0.98 |
| 10 | 0 | 3 | 100 | 50.46 | 0.97 |
| 11 | 0 | 1 | 100 | 20.18 | 0.99 |
| 12 | 0 | 1 | 100 | 40.37 | 0.99 |
| 13 | 0 | 2 | 100 | . | 0.98 |

**Supplemental Table 8. Composite LASSO model scores and associated hypoxemia risk ($SpO_2$<90%), sensitivity, specificity, and positive/negative likelihood ratios (validation dataset)**

| Score | Hypoxemia risk | Sensitivity | Specificity | LR+ | LR- |
|---|---|---|---|---|---|
| -2 | 19 | 7 | 80 | 0.37 | 1.15 |
| -1 | 33 | 7 | 65 | 0.21 | 1.42 |
| 0 | 12 | 5 | 88 | 0.41 | 1.08 |
| 1 | 11 | 13 | 90 | 1.22 | 0.97 |
| 2 | 9 | 8 | 91 | 0.98 | 1.00 |
| 3 | 7 | 13 | 93 | 1.81 | 0.94 |
| 4 | 5 | 17 | 96 | 4.12 | 0.86 |
| 5 | 3 | 13 | 97 | 5.10 | 0.89 |
| 6 | 1 | 6 | 100 | 12.61 | 0.95 |
| 7 | 0 | 4 | 100 | 20.18 | 0.96 |
| 8 | 0 | 2 | 100 | 80.73 | 0.98 |
| 9 | 0 | 1 | 100 | 20.18 | 0.99 |
| 10 | 0 | 2 | 100 | 60.55 | 0.98 |
| 11 | 0 | 1 | 100 | . | 0.99 |

**Supplemental Table 9. Model performance and hypoxemia ($SpO_2$ <90%) case rate (validation dataset)**

| | Hypoxemia risk (n/N) | Crude OR (95% CI) | Mean predicted hypoxemia, % | C-statistic (adjusted for Optimism; 95% CI) | Sensitivity | Specificity | PPV | NPV |
|---|---|---|---|---|---|---|---|---|
| IMCI guideline model | | | | | | | | |
| non-Case | 136/3,472 | Ref | 3.9% | 0.581 (0.580; .533,.613) | 24.4% | 91.8% | 12.9% | 96.1% |
| Case | 44/341 | 3.634 (2.535,5.209) | 12.9% | - | | | | |
| Total | 180 / 3813 | - | | - | | | | |
| WHO-composite model | | | | | | | | |
| non-Case | 54/2,882 | Ref | 1.9% | 0.739 (0.740; .690,.790) | 70.0% | 77.8% | 13.5% | 98.1% |
| Case | 126/931 | 8.197 (5.903,11.382) | 13.5% | - | | | | |
| Total | 180 / 3813 | - | | - | | | | |
| Independent LASSO model | | | | | | | | |
| -2 to -1 | 15/1,277 | Ref | 1.2% | 0.791 (0.789; .742,.833) | 52.8% | 91.6% | 23.8% | 97.5% |
| 0 | 23/1,164 | 1.696 (.881,3.266) | 2.0% | - | | | | |
| 1-2 | 47/972 | 4.275 (2.376,7.692) | 4.8% | - | | | | |
| 3-13 | 95/400 | 26.205 (14.988,45.819) | 23.8% | - | | | | |
| Total | 180 / 3813 | | | - | | | | |
| Composite LASSO model | | | | | | | | |
| -1 | 26/1,996 | Ref | 1.3% | 0.781 (0.780; .725,.816) | 59.4% | 85.4% | 16.8% | 97.7% |
| 1 | 9/452 | 1.539 (.716,3.308) | 2.0% | - | | | | |
| 2 | 38/727 | 4.179 (2.519,6.933) | 5.2% | - | | | | |
| 3-11 | 107/638 | 15.268 (9.842,23.686) | 16.8% | - | | | | |
| Total | 180 / 3813 | | | | | | | |

**Supplemental Table 10. Performance of clinical signs for a $SpO_2$<93% during outpatient pediatric care in Bangladesh and Malawi (validation dataset)**

| Population | Clinical sign | Prevalence $SpO_2$<93% N (%) | C-statistic | Sensitivity | Specificity | LR+ | LR- |
|---|---|---|---|---|---|---|---|
| Overall | WHO danger signs | 57 (50.9) | 0.564 | 14 | 98 | 8.892 | 0.871 |
| | Severe acute malnutrition, WAZ<-3 | 29 (11.7) | 0.504 | 7 | 94 | 1.136 | 0.991 |
| | Stridor | 19 (45.2) | 0.521 | 5 | 99 | 7.129 | 0.958 |
| | Unable to feed | 39 (61.9) | 0.546 | 10 | 99 | 13.97 | 0.908 |
| | Lethargy or unconscious | 8 (47.1) | 0.509 | 2 | 100 | 7.715 | 0.982 |
| | Convulsions | 8 (50) | 0.509 | 2 | 100 | 8.616 | 0.982 |
| | Severe respiratory distress with severe fast breathing | 203 (28.1) | 0.679 | 51 | 85 | 3.356 | 0.578 |
| | Grunting | 62 (49.2) | 0.569 | 16 | 98 | 8.389 | 0.859 |
| | Nasal flaring | 137 (37.6) | 0.64 | 35 | 93 | 5.202 | 0.701 |
| | Head nodding | 69 (35.2) | 0.569 | 18 | 96 | 4.704 | 0.857 |
| | Fever, temp>=38c | 168 (15.8) | 0.592 | 45 | 73 | 1.689 | 0.749 |
| | Severe fast breathing: ≥70 3-11mo; ≥60 12-35mo | 69 (22.5) | 0.553 | 18 | 93 | 2.539 | 0.885 |
| | Central cyanosis (lips/tongue) | 25 (80.6) | 0.531 | 6 | 100 | 36.235 | 0.938 |
| | Chest indrawing | 244 (20.5) | 0.668 | 61 | 72 | 2.212 | 0.534 |
| | Wheezing | 65 (45.5) | 0.57 | 16 | 98 | 7.15 | 0.856 |
| 3-11 months | WHO danger signs | 32 (53.3) | 0.567 | 15 | 98 | 8.88 | 0.864 |
| | Severe acute malnutrition, WAZ<-3 | 15 (18.8) | 0.516 | 7 | 96 | 1.793 | 0.968 |
| | Stridor | 8 (50) | 0.517 | 4 | 100 | 7.838 | 0.967 |
| | Unable to feed | 25 (59.5) | 0.554 | 12 | 99 | 11.419 | 0.892 |
| | Lethargy or unconscious | 4 (57.1) | 0.509 | 2 | 100 | 10.596 | 0.983 |
| | Convulsions | 4 (50) | 0.508 | 2 | 100 | 7.797 | 0.984 |
| | Severe respiratory distress with severe fast breathing | 112 (32) | 0.691 | 53 | 86 | 3.656 | 0.554 |
| | Grunting | 30 (44.8) | 0.56 | 14 | 98 | 6.356 | 0.877 |
| | Nasal flaring | 72 (36.9) | 0.633 | 34 | 93 | 4.564 | 0.713 |
| | Head nodding | 43 (37.1) | 0.58 | 20 | 96 | 4.609 | 0.833 |

| | | | | | | | |
|---|---|---|---|---|---|---|---|
| | Fever, temp>=38c | 82 (17.6) | 0.588 | 41 | 76 | 1.747 | 0.769 |
| | Severe fast breathing: ≥70 3-11mo; ≥60 12-35mo | 22 (26.5) | 0.535 | 11 | 96 | 2.88 | 0.928 |
| | Central cyanosis (lips/tongue) | 14 (82.4) | 0.532 | 7 | 100 | 36.667 | 0.935 |
| | Chest indrawing | 140 (21.4) | 0.673 | 66 | 69 | 2.116 | 0.497 |
| | Wheezing | 26 (42.6) | 0.55 | 12 | 98 | 5.772 | 0.897 |
| 12-23 months | WHO danger signs | 22 (57.9) | 0.575 | 16 | 99 | 11.784 | 0.849 |
| | Severe acute malnutrition, WAZ<-3 | 10 (9.4) | 0.496 | 7 | 92 | 0.893 | 1.01 |
| | Stridor | 10 (52.6) | 0.533 | 7 | 99 | 9.49 | 0.933 |
| | Unable to feed | 12 (66.7) | 0.542 | 9 | 99 | 17.126 | 0.916 |
| | Lethargy or unconscious | 4 (66.7) | 0.514 | 3 | 100 | 17.126 | 0.972 |
| | Convulsions | 3 (50) | 0.51 | 2 | 100 | 8.563 | 0.98 |
| | Severe respiratory distress with severe fast breathing | 67 (26.6) | 0.668 | 50 | 84 | 3.104 | 0.6 |
| | Grunting | 25 (58.1) | 0.586 | 19 | 98 | 12.082 | 0.825 |
| | Nasal flaring | 45 (39.1) | 0.638 | 34 | 94 | 5.551 | 0.707 |
| | Head nodding | 21 (36.2) | 0.562 | 16 | 97 | 4.901 | 0.871 |
| | Fever, temp>=38c | 65 (16.2) | 0.61 | 52 | 70 | 1.747 | 0.687 |
| | Severe fast breathing: ≥70 3-11mo; ≥60 12-35mo | 36 (22.9) | 0.582 | 27 | 90 | 2.564 | 0.817 |
| | Central cyanosis (lips/tongue) | 8 (72.7) | 0.529 | 6 | 100 | 23.178 | 0.942 |
| | Chest indrawing | 75 (20.9) | 0.657 | 56 | 75 | 2.28 | 0.584 |
| | Wheezing | 29 (51.8) | 0.596 | 21 | 98 | 9.205 | 0.804 |
| 24-35 months | WHO danger signs | 3 (21.4) | 0.521 | 6 | 98 | 3.289 | 0.957 |
| | Severe acute malnutrition, WAZ<-3 | 4 (6.5) | 0.492 | 8 | 90 | 0.832 | 1.018 |
| | Stridor | 1 (14.3) | 0.505 | 2 | 99 | 2.044 | 0.989 |
| | Unable to feed | 2 (66.7) | 0.52 | 4 | 100 | 24.612 | 0.961 |
| | Lethargy or unconscious | 0 (0) | 0.497 | 0 | 99 | 0 | 1.007 |
| | Convulsions | 1 (50) | 0.509 | 2 | 100 | 12.306 | 0.981 |
| | Severe respiratory distress with severe fast breathing | 24 (20) | 0.66 | 48 | 84 | 3.015 | 0.618 |
| | Grunting | 7 (43.8) | 0.563 | 14 | 99 | 9.349 | 0.873 |
| | Nasal flaring | 20 (37) | 0.672 | 40 | 94 | 7.094 | 0.636 |
| | Head nodding | 5 (22.7) | 0.537 | 10 | 97 | 3.619 | 0.924 |
| | Fever, temp>=38c | 21 (10.7) | 0.57 | 44 | 70 | 1.475 | 0.8 |
| | Severe fast breathing: ≥70 3-11mo; ≥60 12-35mo | 11 (16.7) | 0.564 | 22 | 91 | 2.408 | 0.858 |

| | | | | | | |
|---|---|---|---|---|---|---|
| Central cyanosis (lips/tongue) | 3 (100) | 0.531 | 6 | 100 | | 0.939 |
| Chest indrawing | 29 (16.1) | 0.665 | 58 | 75 | 2.316 | 0.56 |
| Wheezing | 10 (38.5) | 0.587 | 20 | 97 | 7.538 | 0.822 |

**Supplemental Table 11. Performance of clinical signs for a $SpO_2$<90% during outpatient pediatric care in Bangladesh and Malawi (validation dataset)**

| Population | Clinical sign | Hypoxemia prevalence ($SpO_2$<90%) N (%) | C-statistic | Sensitivity | Specificity | LR+ | LR- |
|---|---|---|---|---|---|---|---|
| Overall | WHO danger signs | 46 (41.1) | 0.622 | 26 | 98 | 14.403 | 0.752 |
| | Severe acute malnutrition, WAZ<-3 | 9 (3.6) | 0.493 | 5 | 93 | 0.778 | 1.016 |
| | Stridor | 15 (35.7) | 0.54 | 9 | 99 | 11.699 | 0.92 |
| | Unable to feed | 34 (54) | 0.593 | 19 | 99 | 24.353 | 0.812 |
| | Lethargy or unconscious | 7 (41.2) | 0.519 | 4 | 100 | 14.692 | 0.962 |
| | Convulsions | 6 (37.5) | 0.516 | 3 | 100 | 12.531 | 0.968 |
| | Severe respiratory distress with severe fast breathing | 111 (15.4) | 0.731 | 63 | 83 | 3.754 | 0.444 |
| | Grunting | 38 (30.2) | 0.597 | 22 | 98 | 9.014 | 0.801 |
| | Nasal flaring | 78 (21.4) | 0.684 | 45 | 92 | 5.663 | 0.602 |
| | Head nodding | 37 (18.9) | 0.585 | 21 | 96 | 4.885 | 0.822 |
| | Fever, temp>=38c | 83 (7.8) | 0.62 | 52 | 73 | 1.877 | 0.668 |
| | Severe fast breathing: ≥70 3-11mo; ≥60 12-35mo | 47 (15.4) | 0.603 | 28 | 93 | 3.876 | 0.779 |
| | Central cyanosis (lips/tongue) | 23 (74.2) | 0.566 | 13 | 100 | 60.659 | 0.868 |
| | Chest indrawing | 125 (10.5) | 0.708 | 71 | 71 | 2.418 | 0.41 |
| | Wheezing | 20 (14) | 0.54 | 11 | 97 | 3.36 | 0.917 |
| 3-11 months | WHO danger signs | 27 (45) | 0.634 | 29 | 98 | 15.441 | 0.726 |
| | Severe acute malnutrition, WAZ<-3 | 4 (5) | 0.5 | 4 | 96 | 0.993 | 1 |
| | Stridor | 7 (43.8) | 0.536 | 8 | 99 | 15.085 | 0.928 |
| | Unable to feed | 22 (52.4) | 0.611 | 23 | 99 | 20.748 | 0.775 |
| | Lethargy or unconscious | 4 (57.1) | 0.521 | 4 | 100 | 25.934 | 0.958 |
| | Convulsions | 2 (25) | 0.509 | 2 | 100 | 6.351 | 0.982 |
| | Severe respiratory distress with severe fast breathing | 61 (17.4) | 0.743 | 65 | 84 | 3.983 | 0.419 |
| | Grunting | 21 (31.3) | 0.6 | 23 | 97 | 8.698 | 0.795 |
| | Nasal flaring | 43 (22.1) | 0.688 | 46 | 91 | 5.39 | 0.588 |
| | Head nodding | 23 (19.8) | 0.599 | 25 | 95 | 4.758 | 0.792 |
| | Fever, temp>=38c | 36 (7.7) | 0.591 | 43 | 75 | 1.737 | 0.759 |
| | Severe fast breathing: ≥70 3-11mo; ≥60 12-35mo | 14 (16.9) | 0.559 | 16 | 96 | 4.037 | 0.877 |

| | | | | | | | |
|---|---|---|---|---|---|---|---|
| | Central cyanosis (lips/tongue) | 14 (82.4) | 0.575 | 15 | 100 | 89.681 | 0.849 |
| | Chest indrawing | 68 (10.4) | 0.697 | 72 | 67 | 2.19 | 0.413 |
| | Wheezing | 6 (9.8) | 0.516 | 6 | 97 | 2.059 | 0.966 |
| 12-23 months | WHO danger signs | 16 (42.1) | 0.618 | 25 | 98 | 14.188 | 0.76 |
| | Severe acute malnutrition, WAZ<-3 | 5 (4.7) | 0.499 | 8 | 92 | 0.966 | 1.003 |
| | Stridor | 7 (36.8) | 0.551 | 11 | 99 | 11.343 | 0.898 |
| | Unable to feed | 10 (55.6) | 0.576 | 16 | 99 | 24.365 | 0.847 |
| | Lethargy or unconscious | 3 (50) | 0.523 | 5 | 100 | 19.492 | 0.955 |
| | Convulsions | 3 (50) | 0.523 | 5 | 100 | 19.492 | 0.955 |
| | Severe respiratory distress with severe fast breathing | 37 (14.7) | 0.706 | 59 | 83 | 3.357 | 0.5 |
| | Grunting | 14 (32.6) | 0.601 | 23 | 98 | 9.562 | 0.793 |
| | Nasal flaring | 23 (20) | 0.645 | 37 | 93 | 4.873 | 0.686 |
| | Head nodding | 12 (20.7) | 0.577 | 19 | 96 | 5.085 | 0.841 |
| | Fever, temp>=38c | 34 (8.5) | 0.636 | 58 | 70 | 1.892 | 0.609 |
| | Severe fast breathing: $\geq$70 3-11mo; $\geq$60 12-35mo | 26 (16.6) | 0.656 | 42 | 89 | 3.928 | 0.65 |
| | Central cyanosis (lips/tongue) | 8 (72.7) | 0.563 | 13 | 100 | 52.774 | 0.873 |
| | Chest indrawing | 44 (12.3) | 0.721 | 70 | 74 | 2.723 | 0.406 |
| | Wheezing | 12 (21.4) | 0.577 | 19 | 96 | 5.32 | 0.84 |
| 24-35 months | WHO danger signs | 3 (21.4) | 0.57 | 16 | 98 | 9.1 | 0.857 |
| | Severe acute malnutrition, WAZ<-3 | 0 (0) | 0.451 | 0 | 90 | 0 | 1.108 |
| | Stridor | 1 (14.3) | 0.523 | 6 | 99 | 5.852 | 0.953 |
| | Unable to feed | 2 (66.7) | 0.555 | 11 | 100 | 70.444 | 0.89 |
| | Lethargy or unconscious | 0 (0) | 0.497 | 0 | 99 | 0 | 1.006 |
| | Convulsions | 1 (50) | 0.527 | 6 | 100 | 35.222 | 0.946 |
| | Severe respiratory distress with severe fast breathing | 13 (10.8) | 0.758 | 68 | 83 | 4.054 | 0.38 |
| | Grunting | 3 (18.8) | 0.569 | 16 | 98 | 7.676 | 0.86 |
| | Nasal flaring | 12 (22.2) | 0.783 | 63 | 93 | 9.534 | 0.395 |
| | Head nodding | 2 (9.1) | 0.54 | 11 | 97 | 3.522 | 0.918 |
| | Fever, temp>=38c | 13 (6.6) | 0.714 | 72 | 70 | 2.447 | 0.394 |
| | Severe fast breathing: $\geq$70 3-11mo; $\geq$60 12-35mo | 7 (10.6) | 0.638 | 37 | 91 | 3.953 | 0.696 |
| | Central cyanosis (lips/tongue) | 1 (33.3) | 0.526 | 6 | 100 | 17.611 | 0.947 |
| | Chest indrawing | 13 (7.2) | 0.71 | 68 | 74 | 2.598 | 0.429 |
| | Wheezing | 2 (7.7) | 0.534 | 11 | 96 | 2.781 | 0.930 |
